# SARS-CoV-2 and the role of vertical transmission from infected pregnant women to their fetuses: systematic review

**DOI:** 10.1101/2021.06.30.21259750

**Authors:** Annette Plüddemann, Elizabeth A. Spencer, Carl J. Heneghan, Jon Brassey, Igho J. Onakpoya, Elena Cecilia Rosca, David H. Evans, John M. Conly, Tom Jefferson

**Affiliations:** University of Oxford, Centre for Evidence-Based Medicine, Nuffield Department of Primary Care Health Sciences, Oxford UK OX2 6GG; Trip Database Ltd, Glasllwch Lane, Newport, UK NP20 3PS; Department of Continuing Education, University of Oxford, Rewley House, 1 Wellington Square, Oxford OX1 2JA, UK; Victor Babes University of Medicine and Pharmacy, Piaţa Eftimie Murgu 2, Timişoara 300041, Romania; Li Ka Shing Institute of Virology and Dept. of Medical Microbiology & Immunology, University of Alberta; Departments of Medicine, Microbiology, Immunology & Infectious Diseases, and Pathology & Laboratory Medicine, Synder Institute for Chronic Diseases and O’Brien Institute for Public Health, Cumming School of Medicine, University of Calgary and Alberta Health Services, Calgary, Canada

## Abstract

**Background:** Vertical transmission of SARS-CoV-2 has been reported but does not appear to be common. This study aims to systematically review the evidence for vertical transmission of SARS-CoV-2.

**Methods:** This review is part of an Open Evidence Review on the transmission dynamics of SARS-CoV-2 and the role of intrauterine mother to fetus transmission. Literature searches were performed in the WHO Covid-19 Database, LitCovid, medRxiv, and Google Scholar for SARS-CoV-2 using keywords and associated synonyms, search date up to 20 December 2020, no language restrictions.

**Results:** We included 106 studies assessing vertical transmission of SARS-CoV-2 from pregnant women to their neonates: these studies comprised 40 reviews (21 fulfilled systematic review methodology, including risk of bias assessment of included studies) and 66 primary studies including 32 case reports (of up to two cases) and 34 prospective and retrospective cohort studies, prospective and retrospective case series, observational studies (including asymptomatic screening), database studies and a quality improvement project. Almost all were conducted in a hospital setting. The 32 case reports were considered to be at high risk of bias, due to the study design; across the 34 remaining primary studies, risk of bias was low to moderate. Sixteen case reports examined vertical transmission, which was not related to maternal symptomatology. For the cohort and case series studies, the percentage of positive neonates ranged from 0% to 22% across the studies. Twenty studies reported no positive vertical transmission. Three studies that reported the highest positivity rates of 11%, 15% and 22% had specifically selected neonates with a positive test (within up to 35 days) within the study population and were therefore more selective populations. Across the cohort and case series studies there were 65/2391 (2.7%) neonates born to mothers with a diagnosis of COVID-19 tested positive for SARS-CoV-2 within 24 hours of birth. No evidence correlated maternal symptomatology to vertical transmission. Mode of delivery did not correlate with rates of vertical transmission. Of 25 studies, 7 identified SARS-CoV-2 in placental tissue; some of these did not demonstrate vertical transmission to the neonate. No study reported the results of viral culture to detect SARS-CoV-2.

**Conclusions:** The results of these studies indicate that vertical transmission is possible, but is not frequent, and factors that influence when vertical transmission occurs are unknown. Further studies using standardised methods to establish viral infection are needed to establish vertical transmission rates and to assess clinical and other conditions affecting transmission.

## Background

Research on previous coronavirus outbreaks of the Middle East respiratory syndrome coronavirus (MERS-CoV) in 2012 and the severe acute respiratory syndrome coronavirus (SARS-CoV) in 2002-2003, assessed the risk of vertical transmission from infected mothers to their neonates and to date no cases of vertical transmission were reported^1,2^. For SARS-CoV-2, several studies have assessed vertical transmission with some early reviews reporting no evidence of vertical transmission ^3,4^, while others reported that vertical transmission was possible and could not be ruled out^4,5^ although the risk appeared to be low ^6^. This study therefore aims to systematically review the evidence for vertical transmission of SARS-CoV-2.

A WHO scientific brief ^2^, published in February 2021, based on an evidence synthesis and expert consultation, considered three mechanisms of vertical transmission: (1) *in utero*, where the virus is present in the blood and crosses the maternal-placental interface; (2) intrapartum, occuring during labour and childbirth via contact with maternal blood, vaginal secretions or faeces; and (3) postnatal, via breast milk but can include contact with an infected mother, another infected caregiver or fomites. The WHO review highlighted that defining vertical transmission of SARS-CoV-2 has been challenging, as much of the reported vertical transmission has been based on a single positive neonatal RT-PCR in an upper respiratory tract specimen, with significant variation in the timing of sample collection. A proposed definition of confirmed *in utero* transmission was considered to be a positive RT-PCR test in one or more neonatal samples (including neonatal blood, respiratory swab, stool sample), amniotic fluid or a placental tissue sample; or positive serology, at age <24 hours, where there was evidence of maternal SARS-CoV-2 infection during pregnancy. Another study^7^ proposed a definition for vertical transmission as including a positive SARS-CoV-2 test in neonates in the first 24h of life (including respiratory tract swab, neonatal blood, cord blood or amniotic fluid) from mothers who had tested positive for SARS-CoV-2 between 14 days prior to birth and 2 days after birth. The UK Obstetric Surveillance System (UKOSS), however, defined vertical transmission as a positive neonatal sample taken within the first 12 h following birth to a mother with confirmed SARS-CoV-2 infection^8^. Given the challenges in defining vertical transmission and that many studies do not report what criteria were applied to define vertical transmission, in the current review, vertical transmission was considered as a positive test in the neonate (including RT-PCR testing of respiratory tract swabs, blood samples, placenta, amniotic fluid) at birth or up to 24h of life, where the mother had either tested positive for SARS-CoV-2 or had a recorded diagnosis of COVID-19.

#### Definition

Vertical transmission: positive test for SARS-CoV-2 in a neonate up to 24 hours of life, including RT-PCR testing of respiratory tract swab samples, fecal samples, blood samples, placenta tissue, amniotic fluid samples, where the mother had a positive test for SARS-CoV-2 or a recorded diagnosis of COVID-19

## Methods

We are undertaking an open evidence review investigating factors and circumstances that impact on the transmission of SARS-CoV-2, based on our published protocol last updated on 1 December 2020 (Supplementary files, Protocol). This review aims to identify, appraise, and summarize the evidence (from peer-reviewed studies or studies awaiting peer review) assessing the occurrence of vertical transmission of SARS-CoV-2 from mothers to their babies in utero. We are conducting an ongoing literature search in the WHO Covid-19 Database, LitCovid, medRxiv, and Google Scholar for

SARS-CoV-2 for keywords and associated synonyms. The search for this review was conducted up to 20 December 2020. We did not impose any language restrictions. (See Supplementary files for the search strategies). Studies of any design (including reviews and case reports) reporting vertical transmission were included. For the included primary studies, the risk of bias was assessed using five domains from the QUADAS-2 criteria ^9^; we adapted this tool because the included studies were not primarily designed as diagnostic accuracy studies. Due to the intrinsic difference in study designs, we did not conduct a quality appraisal of case reports reporting on only one or up to two cases, and data from these studies is reported separately in the Results. We checked the methodology of included reviews and recorded whether they fulfilled systematic review methodology; we did not perform formal assessments of the quality of included systematic reviews but summarized their findings, including quality of their included studies as reported by the authors. We extracted the following information from included primary studies: the country, setting and included population; study design; symptoms reported in mothers (including timing of symptoms if reported); the mode of delivery of babies (if reported); sample sources for SARS-CoV-2 testing for mothers and babies and SARS-CoV-2; and the numbers of neonates testing positive for SARS-CoV-2 from mothers with a COVID-19 positive diagnosis, as well as live culture results and any other relevant results (e.g. assessment of the placenta using histopathology and /or culture and/or immunostaining). We also extracted information on methodology used for RT-PCR, cycle threshold (Ct), and viral culture if reported. We defined “vertical transmission” as a positive test in the neonate within 24h of life, unless otherwise stated.

One reviewer (EAS) assessed the risk of bias from the primary studies (cohort and case series with > 2 cases) and these judgments were independently verified by a second reviewer (AP). Two reviewers (AP and EAS) extracted data from the included primary studies (including the case reports), each extracting data from half of the studes and independently verifying the data extraction of the other studies. One reviewer (AP) extracted data from the included systematic reviews, and these were independently checked by a second reviewer (EAS). Disagreements in the data extraction or bias assessments were resolved by consensus. A third author (IJO) was available in case consensus could not be reached. Results are presented in tabular format and a bar chart was used to represent the percentage of neonates testing positive for SARS-CoV-2 within 24h of birth: these were neonates born to mothers with a SARS-CoV-2 positive test or with a recorded COVID-19 diagnosis which included imaging or X-ray. We did not conduct meta-analysis because of substantial heterogeneity across the included studies.

## Results

We found 123 studies assessing vertical transmission of SARS-CoV-2 from pregnant women to their neonates (Figure 1. PRISMA flow diagram). The studies assessed included 40 reviews and 83 primary studies. Laboratory studies, studies on breast milk only, one study describing a spa outbreak that did not report data for pregnant women, two case reports that did not provide data for testing of the baby, one study on oocytes, and one study that reported a subset of data of another included study^10^ were excluded (see Supplementary files, List of excluded studies). After removing duplicates we included a total of 106 studies: 40 reviews and 66 primary studies.

**Figure 1.**
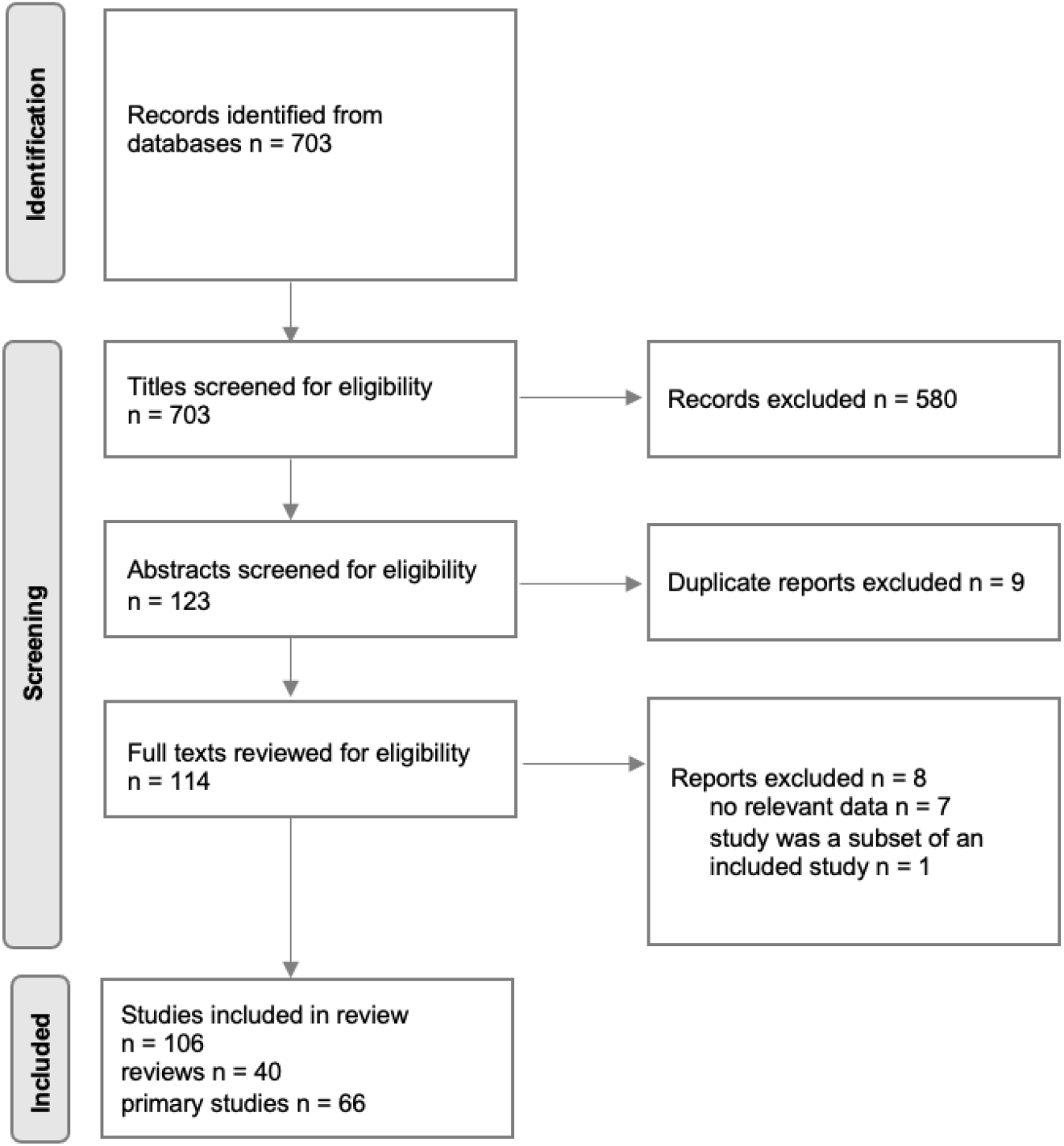
PRISMA flow diagram

Amongst the 66 primary studies or reports (see Supplementary files, Table 1a. Characteristics of included studies), 32 were case reports (describing either single cases only or up to 2 cases) and the remaining 34 studies included prospective and retrospective cohort studies, prospective and retrospective case series, observational studies (including asymptomatic screening), database studies and a quality improvement project. Most studies reported results from testing of neonates, however one cohort study (Smithgall MC 2020) and one case series (Zhang P 2020) reported only on placental samples. Forty eight primary studies reported testing respiratory samples in neonates; in 10 studies the neonatal sample source was not clearly reported; the remainder of the studies examined placental, amniotic, umbilical and skin swab samples. The main findings of the included primary studies and systematic reviews are shown in Supplementary files Table 1b. Main findings of included primary studies and Table 1c. Main findings of included reviews.

### Quality of the evidence

We assessed the methodology of included reviews and documented whether they fulfilled systematic review methodology; we did not otherwise assess their quality. Of 40 reviews, 21 fulfilled systematic review methodology, including risk of bias assessment of included studies; the others are considered narrative reviews. We did not assess the quality of case reports and we did not formally assess the quality of the reviews. The risk of bias of the included primary studies is shown in Table 2. Quality of included primary studies. Almost all studies adequately reported the methods to allow replication of the study with only 5 studies rated as unclear in this domain (although it is to be noted that laboratory testing methods reporting was significantly limited as outlined below). One study did not report information on sample sources and six studies were rated as unclear in this domain, with 5/6 of these studies not reporting the sample sources used for SARS-CoV-2 testing from the mothers, and one not reporting the sample source for the neonates. Analysis and reporting was appropriate for almost all studies with only 2/34 rating as unclear in this domain. Bias was not addressed in 27/34 studies and only 2 studies specifically addressed bias, with the remainder having an unclear rating. We judged the overall risk of bias across the 32 case reports as high; the overall risk of bias across the remaining 34 studies was low to moderate. It is also notable that almost all studies were conducted in a hospital setting and thus only women attending hospital were included in these studies, which may introduce bias regarding the included population in terms of presenting for rapid and accessible SARS-CoV-2 testing, as well as understanding vertical transmission in the out-of-hospital setting.

**Table 2.** Quality assessment of included primary studies.

| **Study** | **Description of methods and sufficient detail to replicate** | **Sample sources clear** | **Analysis & reporting appropriate** | **Is bias addressed** | **Applicability** |
| --- | --- | --- | --- | --- | --- |
| Anand P 2020 | Yes | Yes | Yes | Yes | Yes |
| Ayed A 2020 | Yes | Yes | Yes | Unclear | Yes |
| Bachani S 2020 | Yes | Yes | Yes | Unclear | Yes |
| Barbero P 2020 | Yes | Yes | Yes | Unclear | Yes |
| Chaudhary S 2020 | Yes | Unclear | Yes | No | Yes |
| Cojocaru L 2020 | Yes | Yes | Yes | No | Yes |
| Fenzia B 2020 | Yes | Yes | Yes | No | Yes |
| Gale C 2020 | Yes | Yes | Unclear | Unclear | Unclear |
| Gao X 2020 | Unclear | Yes | Yes | No | Unclear |
| He Z 2020 | Yes | Yes | Yes | No | Yes |
| Hu X 2020 | Yes | Yes | Yes | No | Yes |
| Khan MA 2020 | Yes | Unclear | Yes | No | Yes |
| Liu W 2020 | Yes | Yes | Yes | No | Yes |
| Luo Q 2020 | Yes | Yes | Yes | No | Yes |
| Maraschini A 2020 | Unclear | Yes | Yes | Unclear | Yes |
| Marin Gabriel MA 2020 | Yes | Yes | Yes | No | Yes |
| Masmejan S 2020 | Yes | Yes | Yes | No | Yes |
| Moreno SC 2020 | Yes | Yes | Yes | No | Yes |
| Nayak AH 2020 | Yes | Unclear | Yes | No | Unclear |
| Ogamba I 2020 | Yes | Yes | Yes | No | Yes |
| Olivini N 2020 | Unclear | Yes | Yes | No | Unclear |
| Oncel MY 2020 | Yes | Yes | Yes | No | Yes |
| Pereira A 2020 | Yes | Unclear | Yes | No | Yes |
| Pissarra S 2020 | Yes | Yes | Yes | No | Yes |
| Popofsky S 2020 | Yes | Yes | Yes | No | Unclear |
| Smithgall MC 2020 | Yes | Yes | Yes | No | Yes |
| Schwartz DA 2020 | Unclear | Yes | Yes | No | Unclear |
| Tang F 2020 | Yes | Unclear | Yes | No | Yes |
| Vinuela MC 2020 | Yes | Unclear | Yes | No | Yes |
| Vouga M 2020 | Yes | Yes | Yes | Yes | Yes |
| Woodworth KR 2020 | Unclear | No | Unclear | No | Unclear |
| Yang R 2020 | Yes | Yes | Yes | No | Yes |
| Zhang L 2020 | Yes | Yes | Yes | No | Yes |
| Zhang P 2020 | Unclear | Yes | Yes | No | Unclear |

### Reviews

We included 40 reviews assessing the possibility of vertical maternal-fetal transmission of SARS-CoV-2 (Supplementary file Table 1c. Main findings of included reviews). Half (21/40) of the reviews applied at least some systematic review methodology, including appraisal of the risk of bias of included studies, while the remaining reviews were narrative reviews or literature overviews. The systematic review with the most recent search date, up to 31 October 2020, included 70 studies with a total of 1457 pregnant women diagnosed with COVID-19 (Amaral W 2020). This review identified 21 studies including 39 (3.7%) newborns who tested positive for SARS-CoV-2, and reported the detection of SARS-CoV-2 RNA in the placenta (n = 13). However, only 6 studies in the review reported positive tests for newborns within 24h (n = 13 cases, <1%). The majority (n = 58) of the included studies in the review were judged by the authors to be of low or very low quality using the GRADE quality of evidence assessment, with only one included cohort study judged to be of high quality. Overall 21 reviews (Amaral W 2020, Bellos I 2020, Bwire GM 2020, Caparros-Gonzalez RA 2020, Cavalcante de Melo 2020, Chi J 2020, Deniz M 2020, Dhir SK 2020, Di Toro F 2020, Dube R 2020, Goh XL 2020, Khalil A 2020, Kotlyar AM 2020, Pettirosso E 2020, Raschetti R 2020, Rodrigues C 2020, Romeo G 2020, Sheth S 2020, Tripella G 2020, Turan O 2020, Walker KF 2020) found evidence of vertical transmission, reporting transmission rates between 1% and 5%. Fifteen reviews did not find any evidence for vertical transmission (using a broader definition of vertical transmission including peribirth and post birth), although some stated vertical transmission could not be ruled out despite the lack of evidence (Abd EW 2020, Akhtar H 2020, AlQahtani MA 2020, Della Gatta AN 2020, Diriba K 2020, Gao Y 2020, Han Y 2020, Hessami K 2020, Huntley BJF 2020, Juan J 2020, Novoa RH 2020, Sampieri CL 2020, Shrestha R 2020, Thomas P 2020, Yoon SH 2020). However, for the majority of these reviews, the search end date was until April 2020 or May 2020, with the latest search date being July 2020 for one non-systematic review (Hessami K 2020). The remaining reviews did not specifically report vertical transmission rates. One review reported data solely on transmission via breast milk (Centeno-Tablante E 2020) and found no evidence of SARS-CoV-2 transmission through breast milk. Two reviews focused specifically on maternal characteristics and clinical outcomes and reported that pregnant women did not appear to be more affected by the respiratory complications of COVID-19, compared with non-pregnant women (Figueiro-Filho EA 2020) and that SARS-CoV-2 infection was not specifically shown to increase the risk of maternal, fetal, and neonatal complications (Salem D 2020), although other included reviews have reported the potential for higher risk of complications, depending on the maternal disease course (Amaral W 2020). One review assessed diagnostic methodologies to determine vertical transmission, finding a lack of consensus on the diagnostic strategy for congenital infection (Mahyuddin AP 2020).

### Case reports

There were 32 case reports included in this review, of which 28 reports were of single cases and 4 reports included 2 cases. Sixteen case reports reported vertical transmission, with babies testing positive for SARS-CoV-2 by RT-PCR within 24h, while 13 did not find vertical transmission (Supplementary file Table 1b. Main findings of included primary studies). Three studies did not report results for testing of neonates: three assessed amniotic fluid samples from SARS-CoV-2 PCR positive women only due to death of the fetus, with 2 reporting the presence of SARS-CoV-2 RNA using PCR in amniotic fluid and placental cell supernatant (Shende P 2020, 1 case; Pulinx B 2020, 2 cases) and one reporting a negative test for SARS-CoV-2 RNA (Rubio Lorente AMR 2020). The symptom status of the mother did not directly correlate with whether the baby tested positive, as some asymptomatic women were reported to have a neonate who tested positive for SARS-CoV-2.

### Primary studies

We included 34 primary studies that were not case reports. The primary studies included eight case series (Chaudhary S 2020, Liu W 2020, Masmejan S 2020, Olivini N 2020, Pereira A 2020, Pissarra S 2020, Zhang L 2020, Zhang P 2020), 21 cohort studies (either prospective or retrospective) (Anand P 2020, Ayed A 2020, Bachani S 2020, Barbero P 2020, Gale C 2020, Fenizia B 2020, He Z 2020, Hu X 2020, Luo Q 2020, Khan MA 2020, Nayak AH 2020, Ogamba I 2020, Oncel MY 2020, Maraschini A 2020, Popofsky S 2020, Schwartz DA 2020, Smithgall MC 2020,Tang F 2020, Vinuela MC 2020, Vouga M 2020,Yang R 2020), 2 retrospective observational studies (Gao X 2020, Moreno SC 2020). and one quality improvement project (Cojocaru L 2020), one longitudinal surveillance study (Woodworth KR 2020), and one multi-centre retrospective chart review (Marin Gabriel MA 2020. Across these primary studies we extracted the data reporting the number of neonates testing positive for SARS-CoV-2 by RT-PCR within 24h of birth (typically from a nasopharyngeal swab), who were born to mothers who either had a positive test for SARS-CoV-2 (RT-PCR or serologic testing) or who were diagnosed as having COVID-19 based on clinical assessment including imaging or chest X-ray. Twenty studies reported no positive neonatal cases for neonates tested within 24h of birth (Table 3. Main findings of included cohort and case series studies, Figure 2). Two studies reporting the higher percentage of SARS-CoV-2 cases amongst neonates (22% and 11%) were cohort studies that had specifically selected only neonates with a positive test within 28 days (Gale C 2020) or within 35 days (Schwartz DA 2020) of birth as the included population, where some but not all had been born to mothers with known SARS-CoV-2 infection. These therefore constituted a selected population. The study reporting 15% positivity for neonates was a small study (n=20) including neonates with a positive SARS-CoV-2 test (antibody or RT-PCR) within 14 days after birth or mothers with a positive COVID-19 test in the third trimester (Tang F 2020). These latter studies therefore also constitute a more selected population. The remaining studies reported percentages of positive neonates between 0.8% and 10.8%. Overall there were a total of 2391 neonates included in the analysis of which 65 (2.7%) had a positive RT-PCR test for SARS-CoV-2 within 24h of birth.

**Table 3.** Main findings of included cohort and case series studies.

| Study | Ppts (n) | No. of women diagnosed with COVID-19* | No. of births | No. of positive neonates /no. of neonates tested from women with COVID-19 diagnosis) | No. of positive neonates (RT-PCR) at birth (testing positive within 24h of birth) | Total number of neonates from women with COVID-19 diagnosis | % neonates positive for SARS-CoV-2 RNA | Clinical outcomes for neonates testing positive |
| --- | --- | --- | --- | --- | --- | --- | --- | --- |
| Ayed A 2020 | 185 | 185 | 41 | 2/41 | 0 | 41 | 0.0% | 1 asymptomatic, 1 developed tachypnoea required humidified high flow nasal cannula for 5 days. No deaths |
| Barbero P 2020 | 91 | 91 | 23 | 0/23 | 0 | 23 | 0.0% | n/a |
| Chaudhary S 2020 | 26 | 26 | 14 | 0/14 | 0 | 14 | 0.0% | n/a |
| Cojocaru L 2020 | 1989 | 86 | 34; 31 tested | 0/31 | 0 | 31 | 0.0% | n/a |
| Gao X 2020 | 14 | 14 | 14 | 0/14 | 0 | 14 | 0.0% | n/a |
| He Z 2020 | 22 | 22 | 22 | 0/22 | 0 | 22 | 0.0% | n/a |
| Hu X 2020 | 6 | 6 | 6 | 0/6 | 0 | 6 | 0.0% | n/a |
| Khan MA 2020 | 66 | 66 | 67 | 0/67 | 0 | 67 | 0.0% | n/a |
| Liu W 2020 | 48 | 15 | 48 | 0/15 | 0 | 15 | 0.0% | n/a |
| Luo Q 2020 | 23 | 14 | 23 | 0/14 | 0 | 14 | 0.0% | n/a |
| Masmejan S 2020 (n=13) | 13 | 13 | 13 | 0/13 | 0 | 13 | 0.0% | n/a |
| Moreno SC 2020 | 19 | 19 | 21 | 0/21 | 0 | 21 | 0.0% | n/a |
| Ogamba I 2020 | 40 | 40 | 25; 20 tested | 0/20 | 0 | 20 | 0.0% | n/a |
| Olivini N 2020 | 5 | 5 | 5 | 4/5 | 0 | 5 | 0.0% | Two of the positive neonates developed symptoms possibly related to SARS -CoV-2, no serious illness, no deaths. |
| Pereira A 2020 | 22 | 22 | 22 | 0/22 | 0 | 22 | 0.0% | n/a |
| Pissarra S 2020 | 10 | 10 | 10 | 0/10 | 0 | 10 | 0.0% | n/a |
| Popofsky S 2020 | 160 | 160 | 160 | 1/160 | 0 | 160 | 0.0% | In the cohort, among the 15 symptomatic neonates, 4 had fever, 11 had respiratory distress, 4 had feeding intolerance, 1 had rhinorrhea, and 7 had hypothermia. Only one tested positive for SARS-CoV-2 |
| Vinuela MC 2020 | 100 | 15 | 100 | 0/15 | 0 | 15 | 0.0% | n/a |
| Yang R 2020 | 11078 | 65 | 11078; 38 neonates from 65 positive women tested | 0/38 | 0 | 38 | 0.0% | n/a |
| Zhang L 2020 | 18 | 18 | 18 | 0/18 | 0 | 18 | 0.0% | n/a |
| Oncel MY 2020 | 125 | 125 | 125; 120 tested | 4/120 | 1 | 120 | 0.8% | 1 asymptomatic, 3 required mechanical ventilation or nasal CPAP; however unclear whether these requirements were due to either SARS-CoV-2 infection or prematurity |
| Nayak AH 2020 | 977 | 141 | 134; 131 tested | 3/131 | 3 | 131 | 2.3% | no evidence of transmission of COVID-19 infection |
| Woodworth KR 2020 | 4442 | 4442 | 4495; 610 tested | 16*/610 (*timing of test not reported) | 16 | 610 | 2.6% | 8 of the infants with positive test results were born preterm (26–35 weeks); all were admitted to a neonatal ICU (NICU) without indications reported. Among the 8 term infants with positive test results, one was admitted to a NICU for fever and receipt of supplemental oxygen, one had no information on NICU admission, and the remaining six were not admitted to a NICU. |
| Vouga M 2020 | 1033 | 926 | 820; 384 tested | 11/384 | 11 | 384 | 2.9% | obstetric and neonatal outcomes did not differ between positive and negative patients; comparing positive women with severe maternal outcomes to positive women with no or mild adverse outcomes, showed a significant increased risk of preterm birth and neonatal admission to the intensive care unit |
| Maraschini A 2020 | 146 | 146 | 149 | 9/149 | 5 | 149 | 3.4% | 3 admitted to NICU, none developed serious illness |
| Bachani S 2020 | 348 | 57 | 56 | 5/56 | 2 | 56 | 3.6% | 4 asymptomatic; 1 symptomatic and received respiratory support for 48 hours. No deaths |
| Marin Gabriel MA 2020 | 185 | 242 | 248; 230 tested | 11/230 | 11 | 230 | 4.8% | none presented with pneumonia or any other clinical feature compatible with SARS-CoV-2 infection |
| Fenizia B 2020 | 31 | 31 | 31 | 2/31 | 2 | 31 | 6.5% | all asymptomatic and healthy |
| Anand P 2020 | 69 | 69 | 65 | 7/65 | 7 | 65 | 10.8% | 6/7 asymptomatic, 1 received respiratory support (CPAP) for 48 h, indication: prematurity. No deaths |
| Gale C 2020 | 66 (66 neonates selected based on positivity) | 17 | 66 | 2/17 | 2 | 17 | 11.8% | all asymptomatic and healthy |
| Tang F 2020 | 20 (20 neonates selected based on positivity) | 20 | 20 | 3/20 | 3 | 20 | 15.0% | Infection in infant presumed based on presence of IgM. Symptoms not reported. |
| Schwartz DA 2020 | 19 (19 neonates selected based on positivity) | 9 | 19 | 2/9 | 2 | 9 | 22.2% | 17/19 neonates showed symptoms, including respiratory distress. Selected series of cases. 2 deaths. |

\*based on testing and/or clinical criteria.

**Figure 2.**
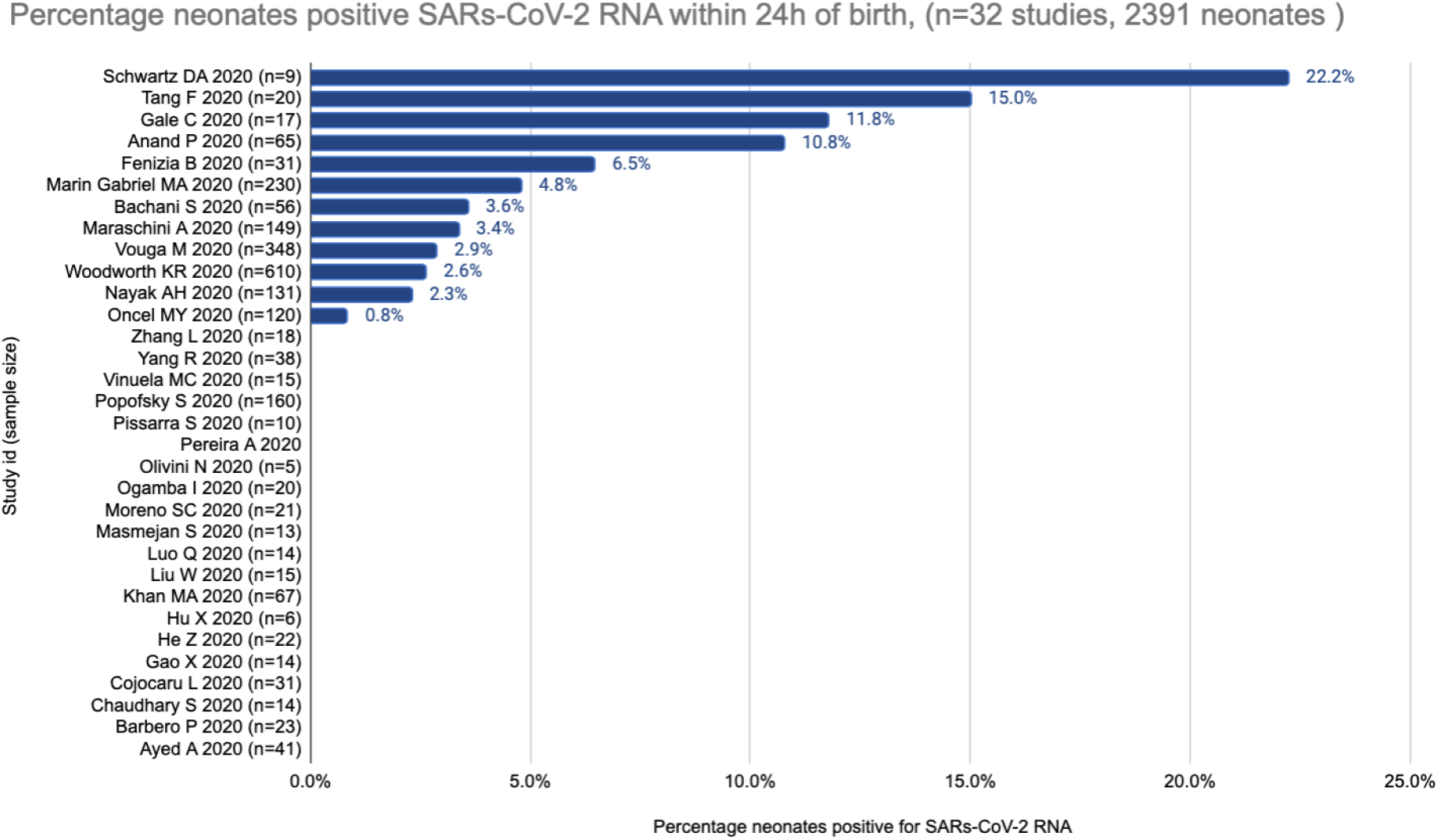
Percentage neonates positive for SARS-CoV-2 within 24h.

The studies included symptomatic and asymptomatic women; however there did not seem to be a specific correlation between symptom status of the mothers and the likelihood of a positive test in the babies, with some studies reporting on women who had symptoms of COVID-19 either immediately prior to or during hospitalisation, some with severe symptoms, where none of the babies tested positive within 24h of birth (e.g. Liu W 2020, Luo Q 2020, Masmejan S 2020, Moreno SC 2020, Ogamba I 2020, Popofsky S 2020) (See Table 1a). Similarly, the mode of delivery (vaginal birth, caesarean section, emergency caesarean section) did not seem to be correlated to positivity rate amongst neonates; across the included cohort and case series studies, for the 6188 cases where mode of delivery was reported, 3939 were by vaginal delivery, 2122 by caesarean section, and 127 reported as emergency caesarean (Table 1b). The overall rate of caesarean section across the included studies thus appears higher than rates generally reported, for example in the UK a recent study reported the rate of caesarean section delivery to range from 13.6% to 31.9% across 146 English NHS trusts^11^, although rates may be context dependent. This may relate to a cautious approach being taken, particularly in the early stages of the pandemic where there was a lack of evidence to inform practice. As noted by others it is reasonable to question whether caesarean delivery for pregnant patients with COVID-19 as the indication, is warranted (Della Gatta AN 2020).

We considered mothers’ test results up to 5 days pre delivery and 8 days post delivery, and mothers’ symptomatology in that same time period (as the relevant time period of likely infectiousness). We also looked at studies’ reports of SARS-CoV-2 testing in the neonates up to or at 24 hours of life, as the most relevant indicator of possible vertical transmission. Testing in mothers was not reported in 4 studies; timing for the mothers’ testing was not reported in 23/79 studies, and the timing was unclear in 1 study. Timing of mothers’ symptomatology was not reported or unclear in 28/83 studies. 75/83 studies reported testing of neonates within 24 hours of life for SARS-CoV-2: among these 75 studies, 144 neonates/1,545 total neonates tested positive within 24h of life. Among those neonates with a positive testing mother within 5 days pre delivery and 8 days post delivery, the number of neonates testing positive was 19/199. For neonates of women with a positive test and symptoms within that time window, 13/32 neonates tested positive within 24h of life. The number of neonates with a positive test within 24hours, born to women with a negative test within 5 days pre and 8 days post delivery, was 4 of 5 neonates.

The focus of this review was to assess the evidence for vertical transmission and did not specifically address the question of maternal and fetal outcomes. Other reviews addressing this question have concluded that SARS-CoV-2 infection was not specifically shown to increase the risk of maternal, fetal, and neonatal complications (Salem D 2020), although some reviews have reported the potential for higher risk of complications, depending on the maternal disease course (Amaral W 2020). The cohort and case series studies in this review reported some cases of neonatal admission to intensive care, but this was not always related to symptoms of SARS-CoV-2 and several studies reported no symptoms in neonates who had tested positive for SARS-CoV-2 (Table 3. Main findings of included cohort and case series studies).

### Detection of virus in the placenta using PCR and immunostaining

Of the included studies, 25 studies reported assessment of the placenta, of these 18 were case studies (Alamar I 2020, Birindwa EK 2020, Ferraiolo A 2020, Grimminck K 2020, Kulkarni R 2020, Hsu AL 2020, Lv Y 2020, Palalioglu RM 2020, Pulinx B 2020, Rebello CM 2020, Shende P 2020, Sisman J 2020, Stonoga E 2020, Tang J 2020, Vivanti AJ 2020, Von Kohorn I 2020, Zaigham M 2020. Zheng T 2020), 2 were retrospective case series (Liu W 2020, Masmejan S 2020), 3 were cohort studies (Fenizia B 2020, Ogamba I 2020, Oncel MY 2020) and 2 were a cohort and a case series specifically studying placental samples (Smithgall MC 2020, Zhang P 2020) (Supplementary file Table Findings of studies reporting placenta analysis). Seven studies reported placentas testing positive for SARS-CoV-2 by in situ hybridisation or immunohistochemistry (Alamar I 2020, Hsu AL 2020, Pulinx B 2020, Shende P 2020, Vivanti AJ 2020, Zaigham M 2020, Zhang P 2020), although not all of these reported vertical transmission to the neonate. Some studies only tested placentas for the presence of SARS-CoV-2 by RT-PCR and did not perform histopathology, while others assessed general inflammation of the placenta, but did not conduct immunostaining for viral presence (Supplementary file Table 4. Findings of studies reporting placenta analysis). One case study conducted whole genome sequencing of viral samples from the mother, placenta and neonate and reported the presence of a single variant of the virus in the mother and placenta. Interestingly, the neonate displayed a SARS-CoV-2 population which included the strain identical to that identified in the mother and placenta, as well as a population with one single-nucleotide polymorphism difference, and they are probably the same strain. This suggests vertical transmission for this case and the potential for a minor intrapatient genetic drift (Zaigham M 2020).

### SARS-CoV-2 testing methodology

None of the included studies reported results of viral culture to detect SARS-CoV-2 (Supplementary file Table 5 Ct counts and RNA concentration results of included studies). All studies used RT-PCR (alone or in combination with clinical and/or immunologic tests) to determine the presence of SARS-CoV-2 infection in the mothers and the babies. However, the majority of studies did not report details of the methods used nor did they report Ct values. Fourteen studies reported RT-PCR methodological information, including the gene probes used (Anand P 2020, Bachani S 2020, Demirjian A 2020, Fenizia B 2020, Gao X 2020, Hinojosa-Velasco A 2020, Luo Q 2020, Pulinx B 2020, Shende P 2020, Tang F 2020, Vinuela MC 2020, Vivanti AJ 2020, Von Kohorn I 2020, Zaigham M 2020), although only 9 studies reported the cycle threshold (Ct) with reported Ct ranging from 13-41 (Bachani S 2020, Demirjian A 2020, Hinojosa-Velasco A 2020, Pulinx B 2020, Shende P 2020, Vinuela MC 2020, Vivanti AJ 2020, Von Kohorn I 2020, Zaigham M 2020). Two studies provided an estimate of viral load based on RT-PCR Ct (Bachani S 2020, Hinojosa-Velasco A 2020), citing a Ct of 15 and 23, respectively, as “high viral load”, and one study reported the viral load of each sample, expressed as log copies/mL or per million of cells as appropriate (Vivanti AJ 2020).

## Discussion

We identified 40 reviews and 66 primary studies assessing vertical transmission of SARS-CoV-2 from pregnant women to their neonates. Overall, the results of these studies indicate that vertical transmission is possible, but is not frequent, and factors affecting whether vertical transmission occurs are unknown. The analysis of the included primary studies that were not case reports found that overall of the 2391 included neonates, 65 (2.7%) had a positive RT-PCR test for SARS-CoV-2 within 24h of birth; this may not be a representative population of pregnant women and neonates. Other systematic reviews included in this study have reported similar results, for example Amaral W 2020 reported a positivity rate for newborns within 24h of less than 1%, Cavalcante de Melo 2020 reported 2% positivity rate and Dhir SK 2020 reported 5% positivity rate. When considering other viruses or microbial pathogens known to be transmitted from mothers to infants, a systematic review of mother-to child transmission of HIV including 18 studies and 6253 participants reported an estimated pooled prevalence of mother-to-child transmission of HIV of 11.4% (95% CI = 9.1-13.7)^12^; a systematic review of vertical transmission of chikungunya virus including 42 studies and 266 infected neonates reported a vertical transmission rate of 50%^13^; and a retrospective analysis of mid-trimester amniocentesis performed in pregnancies with diagnosed maternal infection by cytomegalovirus (CMV), rubella or *Toxoplasma gondii* reported a transmission rate of 17.3% in cases with infection from CMV, 9.5% from *Toxoplasma gondii* and 7.8% from rubella^14^.

All of the included studies in this review used RT-PCR (alone or in combination with clinical and/or immunologic tests) to determine the presence of SARS-CoV-2 infection in the mothers and neonates, but most studies did not report details of the PCR methods used nor did they report Ct values. Notably none of the studies reported results of viral culture to detect SARS-CoV-2 and only one case report conducted whole genome sequencing of viral samples from the mother and neonate, reporting the presence of a single variant of the virus, indicating vertical transmission (Zaigham M 2020). In addition, in most studies no definition was provided as to what constituted “vertical transmission” and the maternal disease course was not always described in relation to testing, therefore it is possible that some cases identified as vertical transmission in the included studies could have been attributable to an alternative source of infection.

The majority of studies utilised PCR testing to assess vertical transmission. Whilst these studies suggest that vertical transmission is possible and may occur in a minority of pregnancies, these data in themselves can not discern whether infection takes place in utero, during the birth process, or via contact with the mother or health care workers at or shortly after birth.

This review did not specifically assess maternal and fetal outcomes. Some early reviews that have assessed this question found that SARS-CoV-2 infection was not specifically shown to increase the risk of maternal, fetal, and neonatal complications (Salem D 2020), while others have reported the potential for higher risk of complications for both mothers and their neonates, depending on the maternal disease course (Amaral W 2020). One recent large multinational cohort study assessing maternal and neonatal morbidity and mortality, comparing pregnant women with and without a COVID-19 diagnosis, found higher rates of adverse outcomes, including maternal mortality, preeclampsia, and preterm birth, in women with COVID-19 (Villar et al 2021) and this warrants further investigation.

The presence of positive tests in both mother and neonate does not unequivocally demonstrate direct transfer of virus from mother to neonate, nor can it demonstrate the biological process by which virus may be transferred. For neonates testing positive for SARS-CoV-2, the virus may have been transmitted directly from mother to neonate across the placenta, but other infection routes may also occur. For example, one study examining fecal samples of mother-neonate dyads found that although the majority of the bacterial microbiome mirrored that of the mother, only a small proportion of the virus population found in a neonate’s fecal samples matched that of the mother. This would be expected given that the gut flora in neonates will be very different from the mother, but this also suggests virus transfer via other routes such as skin contact, breastmilk, or local contamination (Maqsood et al 2019).

Evidence of SARS-CoV-2 RNA in placental samples does not demonstrate placental infection by the virus, nor that transmission via the placental barrier has occurred or can potentially occur (Wastnedge et al 2020). In these studies, positive PCR for SARS-CoV-2 in placentas did not necessarily correlate with positive PCR tests of the neonate. Some studies however showed intervillositis in the placenta and positive immunostaining for SARS-CoV-2 (Pulinx B 2020, Shende P 2020, Vivanti AJ 2020, Zaigham M 2020), which suggests the presence of SARS-CoV-2 in the placenta, and recent work has reported cultivatable SARS-CoV-2 from placental tissue with positive histopathology and whole genome sequencing supporting true vertical transmission (Vayalumkal et al 2021). Overall, viral invasion of the placenta appears to be likely, given the reports in some studies of intervillositis in the placenta and positive immunostaining for SARS-CoV-2; however more high quality information is needed for a clearer understanding of placental infection, the possibility of the virus being transmitted via the placental barrier, and the risk of transmission at the different stages of pregnancy. Nonetheless the demonstration in multiple studies of intervillositis in the placenta and positive immunostaining for SARS-CoV-2 would be strong enough evidence to suggest policy development against placentophagy in the setting of a mother who delivers and has active SARS-CoV-2 infection.

### Strengths and limitations of the study

We conducted a comprehensive search of the literature on vertical transmission of SARS-CoV-2, also including studies that had not yet undergone peer review. We also accounted for the reporting quality of included primary studies. However, we may have missed some relevant studies and recognise that several studies may have been published after December 2020, which was the end date for the search. For example, there may now be more high quality cohort studies as much of the early evidence, as identified in this review, constituted single case studies. The aim of this review was to assess the evidence for vertical transmission and we did not specifically address maternal and fetal outcomes as a primary outcome measure. We did not systematically search for studies reporting this in our review and any conclusions around maternal and fetal outcomes should be interpreted with caution. The definition of vertical transmission varied across studies, with many not reporting the definition of vertical transmission used in the study. It should also be noted that all studies were conducted in the hospital setting, therefore this review does not provide evidence regarding transmission in the out-of-hospital setting.

It is also well known that sample contamination is a hazard of performing PCR testing which is sufficiently sensitive to detect very low concentrations of nucleic acids; for example, to avoid contamination, a study of vertical transmission of HPV used separate buildings for collecting and testing samples (Smith et al 2010). To reduce uncertainty about transmission based on evidence from PCR testing alone, steps for avoiding contamination should be clearly performed and reported. In the studies reviewed here, we found little data relating to PCR procedures and so are unable to evaluate any potential risk of contamination.

## Implications for further research

Evidence of replicable virus as indicated by serial culture with confirmed virus identification, along with concordance in the results of whole genome sequencing for samples from mother and neonate, would substantially reduce uncertainty about the mode of transmission^15^.

Whilst this currently available evidence suggests that vertical transmission is possible and appears to take place infrequently, there is very little information to explain what affects transmission, and therefore how risk can best be mitigated. The risk of vertical transmission is also uncertain when the SARS-CoV-2 infection occurs in the first or second trimester (especially if the mother is asymptomatic). Prospective studies are needed to recruit pregnant women and follow the perinatal and neonatal time course, collecting clinical and exposure information at multiple stages and utilising standardised methods to identify viral infection.

## Conclusion

In conclusion, this review found evidence that vertical transmission can occur, but does not happen frequently. Among neonates born to women with a SARS-CoV-2 positive test or a recorded COVID-19 diagnosis, the great majority do not test positive for SARS-CoV-2 within the first 24 hours of life, indicating a low rate of vertical transmission. From the data we examined, it is not possible to establish what factors may affect vertical transmission.

## Supporting information

PRISMA checklist

Protocol

List of excluded studies

Literature search strategy

## Data Availability

This systematic review uses only data published in scientific journals or on preprint websites and thus in the public doman.

## Competing Interests

TJ was in receipt of a Cochrane Methods Innovations Fund grant to develop guidance on the use of regulatory data in Cochrane reviews (2015-018). In 2014–2016, he was a member of three advisory boards for Boehringer Ingelheim. TJ was a member of an independent data monitoring committee for a Sanofi Pasteur clinical trial on an influenza vaccine. TJ is occasionally interviewed by market research companies about phase I or II pharmaceutical products for which he receives fees (current). TJ was a member of three advisory boards for Boehringer Ingelheim (2014-16). TJ was a member of an independent data monitoring committee for a Sanofi Pasteur clinical trial on an influenza vaccine (2015-2017). TJ is a relator in a False Claims Act lawsuit on behalf of the United States that involves sales of Tamiflu for pandemic stockpiling. If resolved in the United States favor, he would be entitled to a percentage of the recovery. TJ is coholder of a Laura and John Arnold Foundation grant for development of a RIAT support centre (2017-2020) and Jean Monnet Network Grant, 2017-2020 for The Jean Monnet Health Law and Policy Network. TJ is an unpaid collaborator to the project Beyond Transparency in Pharmaceutical Research and Regulation led by Dalhousie University and funded by the Canadian Institutes of Health Research (2018-2022). TJ consulted for Illumina LLC on next generation gene sequencing (2019-2020). TJ was the consultant scientific coordinator for the HTA Medical Technology programme of the Agenzia per i Servizi Sanitari Nazionali (AGENAS) of the Italian MoH (2007-2019). TJ is Director Medical Affairs for BC Solutions, a market access company for medical devices in Europe. TJ was funded by NIHR UK and the World Health Organization (WHO) to update Cochrane review A122, Physical Interventions to interrupt the spread of respiratory viruses. TJ is funded by Oxford University to carry out a living review on the transmission epidemiology of COVID-19. Since 2020, TJ receives fees for articles published by The Spectator and other media outlets. TJ is part of a review group carrying out Living rapid literature review on the modes of transmission of SARS-CoV-2 (WHO Registration 2020/1077093-0). He is a member of the WHO COVID-19 Infection Prevention and Control Research Working Group for which he receives no funds. TJ is funded to co-author rapid reviews on the impact of Covid restrictions by the Collateral Global Organisation. He is also an editor of the Cochrane Acute Respiratory Infections Group. TJ’s competing interests are also online https://restoringtrials.org/competing-interests-tom-jefferson

CH holds grant funding from the NIHR, the NIHR School of Primary Care Research, the NIHR BRC Oxford and the World Health Organization for a series of Living rapid review on the modes of transmission of SARs-CoV-2 reference WHO registration No2020/1077093. He has received financial remuneration from an asbestos case and given legal advice on mesh and hormone pregnancy tests cases. He has received expenses and fees for his media work including occasional payments from BBC Radio 4 Inside Health and The Spectator. He receives expenses for teaching EBM and is also paid for his GP work in NHS out of hours (contract Oxford Health NHS Foundation Trust). He has also received income from the publication of a series of toolkit books and for appraising treatment recommendations in non-NHS settings. He is Director of CEBM, an NIHR Senior Investigator and an advisor to Collateral Global. He is also an editor of the Cochrane Acute Respiratory Infections Group

DHE holds grant funding from the Canadian Institutes for Health Research and Li Ka Shing Institute of Virology relating to the development of Covid-19 vaccines as well as the Canadian Natural Science and Engineering Research Council concerning Covid-19 aerosol transmission. He is a recipient of World Health Organization and Province of Alberta funding which supports the provision of BSL3-based SARS-CoV-2 culture services to regional investigators. He also holds public and private sector contract funding relating to the development of poxvirus-based Covid-19 vaccines, SARS-CoV-2-inactivation technologies, and serum neutralization testing.

JMC holds grants from the Canadian Institutes for Health Research on acute and primary care preparedness for COVID 19 in Alberta, Canada and was the primary local Investigator for a Staphylococcus aureus vaccine study funded by Pfizer for which all funding was provided only to the University of Calgary. He is a co investigator on a WHO funded study using integrated human factors and ethnography approaches to identify and scale innovative IPC guidance implementation supports in primary care with a focus on low resource settings and using drone aerial systems to deliver medical supplies and PPE to remote First Nations communities during the COVID 19 pandemic. He also received support from the Centers for Disease Control and Prevention (CDC) to attend an Infection Control Think Tank Meeting. He is a member of the WHO Infection Prevention and Control Research and Development Expert Group for COVID 19 and the WHO Health Emergencies Programme (WHE) Ad hoc COVID 19 IPC Guidance Development Group, both of which provide multidisciplinary advice to the WHO, for which no funding is received and from which no funding recommendations are made for any WHO contracts or grants. He is also a member of the Cochrane Acute Respiratory Infections Group.

JB is a major shareholder in the Trip Database search engine (www.tripdatabase.com) as well as being an employee. In relation to this work Trip has worked with a large number of organisations over the years, none have any links with this work. The main current projects are with AXA and Collateral Global.

ECR was a member of the European Federation of Neurological Societies(EFNS) / European Academy of Neurology (EAN) Scientist Panel – Subcommittee of Infectious Diseases (2013-2017). Since 2021, she is a member of the International Parkinson and Movement Disorder Society (MDS) Multiple System Atrophy Study Group and the Mild Cognitive Impairment in Parkinson Disease Study Group. She was an External Expert and sometimes Rapporteur for COST proposals (2013, 2016, 2017, 2018, 2019) for Neurology projects.

AP is Senior Research Fellow at the Centre for Evidence-Based Medicine and reports grant funding from NIHR School of Primary Care Research (NIHR SPCR ESWG project 390 and project 461), during the conduct of the study; and occasionally receives expenses for teaching Evidence-Based Medicine. IJO, EAS have no interests to disclose.

## Acknowledgements

This work was commissioned and paid for by the World Health Organization (WHO). Copyright on the original work on which this article is based belongs to WHO. The authors have been given permission to publish this article. The author(s) alone is/are responsible for the views expressed in the publication. They do not necessarily represent views, decisions, or policies of the World Health Organization.

## Data Availability

All data included in the review are provided in the tables or in the supplemental files.

