## Supplementary material for "SARS-CoV-2 and the role of vertical transmission from infected pregnant women to their fetuses: systematic review": Protocol

### The evidence on transmission dynamics of COVID-19: protocol for a series of Systematic Reviews

Protocol for a living evidence review (Version 3: 1 December 2020)

**Affiliations**

1. The University of Oxford.
2. Trip Database Ltd. https://www.tripdatabase.com

**Keywords**

COVID-19; SARS-CoV-2; Transmission.

---

**Background**

COVID-19 is a new disease, distinct from other diseases caused by coronaviruses, such as Severe Acute Respiratory Syndrome (SARS) and Middle East Respiratory Syndrome (MERS). The SARS-CoV-2 virus spreads rapidly. At present, there are no therapeutics or vaccines proven to treat or prevent COVID-19, although national governments, WHO and partners are working urgently to coordinate the response and rapid development of prevention, control and management measures on many fronts.

The overarching aim of the WHO's global Strategic Preparedness and Response Plan for COVID-19 [1] is to control COVID-19 by suppressing transmission of the virus and preventing associated illness and death. However, transmission of the SARS-CoV-2 virus and the disease it causes is poorly understood, and public health measures for restricting transmission are based on limited information with relatively few systematic reviews on the transmission of the SARS-CoV-2 virus available.

Given the novelty of the disease and its cause, early reliance on models of spread is based on what is known of the dynamics of other respiratory infections, especially those due to other coronaviruses and influenza. One of the most important aspects of these uncertainties regards the modes and circumstances of transmission of newly identified agents. As a result, research is ongoing throughout the world across various disciplines with the aim of understanding SARS-CoV-2 modes of transmission, complemented with rapid publications. As a result, there is a need to continuously and systematically conduct reviews of publicly available studies with the latest knowledge from publications to inform WHO recommendations using the most up-to-date information.

**Objectives**

Objectives are to provide a rapid summary and evaluation of relevant data on transmission of SARS-CoV-2, report important policy implications, and highlight areas of research urgently needed. These transmission areas include airborne, contact and droplet, orofecal, vertical, fomite and other modes such as urine and blood and body fluids.

---

[1] Operational planning guidance to support country preparedness and response. Geneva: World Health Organization; 2020 https://www.who.int/publications/i/item/draft-operational-planning-guidance-for-un-country-teams

#### Modes of transmission for SARS-CoV-2 [2] [3]

Respiratory droplets are >5-10 µm in diameter. Respiratory droplets that include virus can reach the mouth, nose or eyes of a susceptible person and can result in infection.

- Respiratory droplets <5µm in diameter are referred to as droplet nuclei or aerosols. Airborne transmission is the spread of an infectious agent caused by the dissemination of aerosols that remain infectious when suspended in air over long distances and time.
- Close or direct contact transmission occurs with an infected person who has respiratory symptoms.
- Respiratory secretions or droplets can contaminate surfaces and objects, creating fomites (contaminated surfaces).
- Orofecal transmission occurs where the virus in fecal particles can pass from one person to the mouth of another. Main causes include lack of adequate sanitation and poor hygiene practices. Fecal contamination of food is another form of orofecal transmission.
- Intrauterine/ mother to child transmission of SARS-CoV-2 from infected pregnant women to their fetuses ( vertical transmission).
- Bloodborne or body fluid transmission.

**Subgroups:**

Where feasible will assess transmission outcomes by setting:healthcare facilities, community settings and the environment. We will report the evidence from studies with the results of reverse transcriptase polymerase chain reaction (RT-PCR) where reported by cycle threshold, time from symptom onset and live culture of SARS-CoV-2 by transmission mode. Evidence from studies comparing culture with other means of diagnosis is not usually mode of transmission-specific. Updates of the culture review [4] will be carried out alongside and in parallel with mode of transmission study extraction for all modes.

#### Methods

*Search Strategy*

The following electronic databases are searched, with searches being updated approximately each month starting from 1 December 2020 with screening every three months unless stated otherwise: LitCovid, medRxiv, Google Scholar and the WHO Covid-19 database. Search terms are Covid-19, SARS-CoV-2, transmission, and appropriate synonyms. The reference lists of included studies are searched for additional relevant studies.

---

##### Search Strategy (last updated 30 Jan 2021)

We searched four main databases:,

- **WHO Covid-19 Database** (https://search.bvsalud.org/global-literature-on-novel-coronavirus-2019-ncov/)

---

[2] Transmission of SARS-CoV-2: implications for infection prevention precautions: https://www.who.int/news-room/commentaries/detail/transmission-of-sars-cov-2-implications-for-infection-prevention-precautions

[3] Infection Prevention and Control of Epidemic-and Pandemic-prone Acute Respiratory Infections in Health Care. Geneva: World Health Organization; 2014 (available at https://apps.who.int/iris/bitstream/handle/10665/112656/9789241507134_eng.pdf;jsessionid=41AA684FB64571CE8D8A453C4F2B2096?sequence=1).

[4] Jefferson T; Spencer EA; Brassey J; Heneghan C. Viral cultures for COVID-19 infectious potential assessment – a systematic review. Clinical Infectious Diseases 2020 (in press).

The global literature cited in the WHO COVID-19 database is updated daily (Monday through Friday) from searches of bibliographic databases, hand searching, and the addition of other expert-referred scientific articles.

- **LitCovid** (https://www.ncbi.nlm.nih.gov/research/coronavirus/)
  A curated literature hub for tracking up-to-date scientific information about the 2019 novel Coronavirus. It is a comprehensive resource on the subject, providing central access to relevant articles in PubMed.
- **medRxiv** (https://www.medrxiv.org/)
  A free online archive and distribution server for complete but unpublished manuscripts (preprints) in the medical, clinical, and related health sciences.
- **Google Scholar** (https://scholar.google.com/)
  Provides a broad search for scholarly literature across many disciplines and sources: articles, theses, books, abstracts and court opinions, from academic publishers, professional societies, online repositories, universities and other web sites.

Searches were initially undertaken monthly but from September, due to the volume of new studies, they moved to search every fortnight. For databases which do not support date granularity, date of publication was approximated. All retrieved articles are entered into a google sheet that is then used to filter and check articles for inclusion.

WHO Covid-19 Database, LitCovid are specific Covid-19 databases so there was no requirement to add a search string to identify Covid-19 articles. For medRxiv and Google Scholar, we used the terms *coronavirus OR covid-19 OR SARS-CoV-2*.

For the respective topics we used the following terms:

Airborne: *aerosol OR airborne OR airbourne OR inhalation OR air OR droplet*
Orofecal: *orofecal OR oro-fecal OR faecal OR fecal OR stool OR faeces OR feces OR rectal OR rectum OR anal OR anus OR toilet*
Fomite: *fomite OR surfaces*

These terms were also combined with the term *transmission*.

Additional techniques were used to identify relevant topics:

- For articles that looked particularly relevant, citation tracking was undertaken.
- For included systematic reviews, the lists of included and excluded articles were examined for inclusion.

The search strategy is subject to continual review and updated and is maintained by Brassey J.

##### *Study inclusion and exclusion*

Eligible studies should include sampling for the detection of SARs-CoV-2 in the population or the environment on any potential mode of transmission, including droplet, airborne, fomite, orofecal, bloodborne, vertical or other. Studies can be observational including case series, ecological, or prospective; or interventional including randomised trials and clinical reports, outbreak reports, case-control studies, experimental studies, non-predictive modelling. Studies should include sampling for the detection of SARs-CoV-2

Studies on factors influencing transmission are included, such as location settings, meteorological or immunological factors. Studies incorporating models to describe observed data are included. Studies reporting solely predictive modelling are excluded.

##### *Data extraction*

Study data are extracted into data extraction templates Table 1 (study characteristics) and Table 2 (methodological quality of studies) and Table 3 (summary of main findings). References are included in alphabetical order as a webappendix that facilitates updating. We follow PRISMA reporting guidelines as indicated for systematic or scoping reviews where applicable (PRISMA checklist) [5] Extraction is performed by one author and checked by a second author. Where there is disagreement, a third author arbitrates.

*Quality assessment*

Included studies quality is assessed based on a modified Quadas-2 tool using five criteria: (1) a clearly defined setting; (2) demographic characteristics or sampling procedures adequately described; 3) follow-up duration sufficient for the outcomes; (4) the transmission outcomes assessed adequately; (5) main biases that are threats to validity taken into consideration. Quality assessment is performed by one author and checked by a second author. In the case of disagreement a third author arbitrates, or for culture

*Data synthesis and reporting*

Outcomes are specified within each review. We summarise data narratively and report the outcomes as stated in the paper, including quantitative estimates where feasible and relevant. We report the detection of a live culture of SARS-CoV-2 when reported (see also *subgroups*). Where possible, compatible datasets may be pooled for meta-analysis. We may write to authors for clarification of data, and also report research and policy implications.

*Continual data release*

Summary descriptions of important relevant research papers identified are summarised in the tracker and corresponding folders in an ongoing manner.  As important new data accumulates, we produce a report as an individual rapid review and aim to make all our work available by depositing the review findings on the Oxford Research Archive. [6]

**Funding**

This work is supported by the National Institute of Health Research Evidence Synthesis Working Group [Project 380] and funded by the World Health Organization Living rapid review on the modes of transmission of SARs-CoV-2 reference WHO registration No2020/1077093.

---

[5] Tricco AC, Lillie E, Zarin W et al. PRISMA extension for scoping reviews (PRISMA-ScR): checklist and explanation. Ann Intern Med. 2018,169(7):467-473. doi:10.7326/M18-0850
[6] The Oxford Research Archive. Website. https://ora.ox.ac.uk Accessed 5 October 2020.
