## Supplementary Tables for "SARS-CoV-2 and the role of vertical transmission from infected pregnant women to their fetuses: systematic review"

### **Table 1a.** Characteristics of included primary studies

| Study ID | Setting | Country | Population/environment | Method |
|---|---|---|---|---|
| Alamar I 2020 | Hospital | USA | 1 mother with fever, mild chills, fatigue, dysgeusia, and anosmia beginning 1 day before presentation, with positive COVID-19 test. | Case study |
| Anand P 2020 | Hospital | India | 69 COVID19 + mothers, 65 infants (1 abortion, 4 stillbirths). 51 mothers had symptoms and of these 45 had mild symptoms. | Cohort study |
| Ayed A 2020 | Hospital | Kuwait | 185 pregnant women with PCR confirmed SARS-CoV-2 infection (median age 31; median gestation at diagnosis 29 weeks). 88% had mild symptoms, (fever & cough the most common presenting symptoms). During the study period 40 (21.6%) gave live birth, 3 (1.6%) had a miscarriage, 1 (0.54%) had intrauterine fetal death, unrelated to COVID19. 2 (1.1%) patients developed severe pneumonia & required intensive care. Most neonates asymptomatic; 2 (5%) of them tested positive on day 5. | Retrospective study, medical records |
| Bachani S 2020 | Hospital | India | 348 pregnant women tested for SARS-CoV-2, 57 women (16.3%) confirmed positive. 3 maternal deaths were associated with comorbidities. 5 neonates tested positive for SARS-CoV-2 | Retrospective study |
| Bandyopadhyay T 2020 | Hospital | India | 1 mother, mild grade fever, close contact of positive case (husband) | Case study |

| | | | | |
|---|---|---|---|---|
| Barbero P 2020 | Hospital | Spain | 91 women with COVID-19 symptoms (fever (37.8 C), dry cough, shortness of breath or dyspnea, chills & myalgia, headache, coryza & new onset of loss of taste or smell) & diagnosed with SARS-CoV-2 infection during pregnancy/postpartum (<40 days after giving birth). 46.2% rate of hospitalization. | Retrospective cohort study |
| Birindwa EK 2020 | Hospital | DR Congo | 1 woman positive for SARS-Cov-2. | Case report |
| Bordbar A 2020 | Hospital | Iran | 1 women, gestational diabetes, no COVID-19 symptoms; tested 2 days after giving birth. | Case report |
| Chaudhary S 2020 | Hospital | Pakistan | 26 women, RT-PCR COVID-19 positive; 10 asymptomatic, 16 symptomatic; all mild disease, 1 pneumonia. | Case series |
| Cojocaru L 2020 | Hospital | US | 1989 women were screened for SARS-CoV-2, from which 86 (0.04%) tested positive. 34 delivered during study period (3 excluded from analysis). 5 women in analysis admitted to ICU. | Quality improvement project, prospective |
| Demirjian A 2020 | Hospital | UK | 1 woman, fever & respiratory symptoms, tested for COVID on day 6 of admission. | Case report |
| Farsi Z 2020 | Hospital | Iran | 1 woman, pregnant with triplets, symptoms of cough, fever, myalgia | Case report |
| Fenizia B 2020 | Hospital | Italy | 31 laboratory-confirmed COVID19+ women, also radiological assessment, signs & symptoms; 4 severe cases requiring ICU admission | Cohort study, multicentre |
| Ferraiolo A 2020 | Hospital | Italy | 1 woman, no symptoms, breech presentation | Case report |
| Gale C 2020 | Hospital; National Registry | UK | Infants with a diagnosis of SARS-CoV-2 within 28 days | Prospective cohort study |
| Gao W 2020 | Hospital | China | 1 asymptomatic woman | Case report |

| | | | | |
|---|---|---|---|---|
| Gao X 2020 | Hospital | China | 14 pregnant women with lab-confirmed COVID-19 | Retrospective observational study |
| Grimminck K 2020 | Hospital | The Netherlands | 1 pregnant woman, oropharyngeal swab RT-PCR positive for COVID-19, immunosuppressed (lupus) | Case report |
| He Z 2020 | Hospital | China | 22 newborns born to pregnant women with COVID-19 | Retrospective cohort study |
| Hinojosa-Velasco A 2020 | Hospital | Mexico | 1 symptomatic woman (fever, cough, sore throat), patchy chest radiograph, testing positive for COVID-19 | Case report |
| Hsu AL 2020 | Hospital | USA | 1 pregnant woman, mild symptoms | Case report |
| Hu X 2020 | Hospital | China | 6 preterm infants born to COVID-19 positive mothers | Retrospective cohort study |
| Khan MA 2020 | Hospital | Pakistan | 66 pregnant women testing COVID-19 positive, most asymptomatic | Cohort study |
| Kulkarni R 2020 | Hospital | India | Pregnant woman, admitted with fever & body ache, in active labor | Case report |
| Liu W 2020 | Hospital | China | Neonates born to women with: 1. confirmed COVID-19 (symptoms & positive RT-PCR), 2. suspected COVID-19 (symptoms with negative RT-PCR but chest CT pneumonia), 3. control cases ( with or without symptoms, confirmed as influenza) | Retrospective analysis of cases |
| Luo Q 2020 | Hospital | China | 23 pregnant women, 14 with confirmed COVID-19, 9 with suspected COVID-19 (chest-CT pneumonia but negative PCR and serology). | Cohort study of breast feeding |
| Lv Y 2020 | Hospital | China | 1 pregnant woman, fever & cough, SARS-CoV-2 PCR positive | Case report |

| | | | | |
|---|---|---|---|---|
| Maraschini A 2020 | Hospital | Italy | 146 pregnant women with confirmed COVID-19, 142 confirmed by RT-PCR, 4 confirmed by chest X-ray; 99 no COVID pneumonia, 47 with COVID pneumonia | Cohort study |
| Marin Gabriel MA 2020 | Hospital | Spain | Pregnant women, third trimester, RT-PCR or serology OCVID-19 positive: 222 RT-PCR positive; 19 RT-PCR negative but serology positive at delivery. | Multi-centre study, retrospective chart review plus 1 month follow-up |
| Masmejan S 2020 | Hospital | Switzerland | 13 pregnant women, positive RT-PCR or positive serology (IgG) for SARS-CoV-2. 11 mild symptoms or asymptomatic, 2 critical fever, severe symptoms | Retrospective case series |
| Mohakud NK 2020 | Hospital | India | Symptomatic pregnant woman, taking treatment for hypothyroidism, at 32+ weeks tested positive for SARS-CoV-2. | Case report |
| Moreno SC 2020 | Hospital maternity unit | USA | Symptomatic pregnant women diagnosed positive for COVID-19 via PCR in the third trimester, & all neonates with complete COVID-19 testing & delivery data. | Retrospective observational study |
| Nayak AH 2020 | Tertiary care hospital maternity unit | India | Pregnant women attending for obstetric care. | Cohort study |
| Ogamba I 2020 | Four acute care hospitals | USA | Pregnant women 18 years or older with a diagnosis of COVID-19 (testing was performed on admission to the labor & delivery unit, outpatient setting, and/or inpatient hospitalization) | Retrospective cohort study |
| Olivini N 2020 | COVID-19 ward of children's hospital | Italy | 5 women who tested positive after screening due to close contact with an infected maternity services HCW, & their neonates. | Retrospective case series |
| Oncel MY 2020 | Hospital maternity units, multicentre | Turkey | 125 pregnant women & their neonates, tested if symptomatic or had had close contact with family members with COVID-19. | Cohort study |

| | | | | |
|---|---|---|---|---|
| Palalioglu RM 2020 | Hospital maternity unit | Turkey | Woman aged 42 years, 37 weeks pregnant, with diet-regulated gestational diabetes mellitus, tested SARS-CoV-2 positive, asymptomatic. Admitted two weeks later & C-section performed due to prelabour rupture of membrane. | Case report |
| Parsa Y 2020 | Hospital maternity unit | Iran | 41-yr-old pregnant woman with signs & symptoms of acute respiratory illness presented with labor pain &vaginal leak at 37 weeks gestation, tested positive for COVID-19 using RT-PCR, emergency C-section. | Case report |
| Pereira A 2020 | Hospital maternity unit | Spain | 22 pregnant women with SARS-CoV-2 (11 symptomatic), median age 34 years, median gestational age 38+5 weeks. | Case series |
| Pessoa FS 2020 | Hospital maternity unit | Brazil | 34 year old woman 33+ weeks pregnant, RT-PCR positive, admitted with flu-like symptoms including fever, dry cough & fatigue from talking; chest computed tomography showed attenuations with ground glass opacification and bilateral consolidations. | Case report |
| Pissarra S 2020 | Hospital maternity unit | Portugal | 10 SARS-CoV-2 positive pregnant women (7 were symptomatic) giving birth at the hospital. | Case series |
| Popofsky S 2020 | Telephone survey | USA | Women with positive SARS-CoV-2 PCR & their neonates, born at one of 3 hospitals in New York State. | Observational longitudinal cohort |
| Pulinx B 2020 | Hospital obstetric department. | Belgium | 30 year old woman 22 weeks pregnant with dichorionic diamniotic twins, recently diagnosed with gestational diabetes mellitus, tested positive for SARS-CoV-2 by RT-PCR. | Case report |
| Rebello CM 2020 | Hospital maternity unit | China | 32-year-old woman pregnant at 34 weeks, with gestational diabetes; symptoms of anosmia, ageusia, runny nose, dry cough, headache, myalgia, no fever, diagnosed with SARS-CoV-2. | Case report |

| | | | | |
|---|---|---|---|---|
| Rivera-Hernandez P 2020 | Hospital | USA | 38-year-old pregnant woman (32/33 weeks) with obesity, poorly controlled type 2 diabetes, asthma & 3-day history of dyspnea & malaise, admitted to ICU. | Case report |
| Rubio Lorente AM 2020 | Hospital | Spain | 2 pregnant women, 1: 20w pregnant, fever and cough, positive SARS-CoV-2 PCR test, negative serology. 2: 12w pregnant, delayed abortion, low fever, serology positive, PCR-negative for SARS-CoV-2 | Analysis of amniotic fluid of 2 cases |
| Sajjan GR 2020 | Hospital maternity unit | India | 21yr old primigravida at 32 weeks gestational age, diagnosed with PPROM | Case report |
| Schwartz DA 2020 | 10 hospitals in different cities in Iran | Iran | 19 neonates testing positive for SARS-CoV-2 | Cohort study |
| Shende P 2020 | Hospital maternity unit | India | 1 woman, asymptomatic, 8 weeks pregnant, tested positive for SARS-CoV-2 by PCR. Fetal demise at 13 weeks followed by dilation & curettage termination procedure. | Case report |
| Singh MV 2020 | Hospital maternity unit | India | 1 asymptomatic pregnant woman with positive SARS-CoV-2 test, & her neonate | Case report |
| Sisman J 2020 | Hospital maternity unit | USA | 1 pregnant woman with obesity, diabetes mellitus & late latent syphilis, & her neonate. | Case report |
| Smithgall MC 2020 | Hospital | USA | 3rd trimester placentas from 51 SARS-CoV-2-positive and 25 SARS-CoV-2-negative women. | Cohort study (placentas) |
| Stonoga E 2020 | Hospital | Brazil | Woman at 27 weeks' gestation with COVID-19 symptoms: dyspnea, dry cough, high temperature (38.5°C), anosmia, nausea, vomiting, & diarrhea had developed 2 days before hospitalization. | Case report |

| | | | | |
|---|---|---|---|---|
| Tang F 2020 | Hospital | China | Neonates born during the pandemic period 2 February to 31 March 2020 in Wuhan, whose mother was confirmed with COVID-19 in the 3rd trimester, or with +ve SARS-CoV-2-IgM and SARS-CoV-2-IgG and/or +ve RT-PCR for SARS-CoV-2 within 14 days after birth. All included neonates had excluded diagnoses of adenovirus, enterovirus, influenza A, influenza B, parainfluenza, chlamydophila pneumonia and mycoplasma pneumonia. | Observational cohort |
| Tang J 2020 | Hospital | China | Two mothers with symptomatic COVID-19 in the second trimester and their neonates. | Case report (2 cases) |
| Vendola N 2020 | Hospital maternity unit | Italy | 2 pregnant women with positive IGG on peripheral blood test. | Case report (2 cases) |
| Vinuela MC 2020 | Hospital maternity unit | Spain | Systematic screening in asymptomatic women admitted for spontaneous delivery: first 100 consecutive participants; 9 tested positive for SARS-CoV-2 by PCR; 13 (including 7 of the PCR-positive women) had antibodies to SARS-CoV-2. | Observational study, asymptomatic screening |
| Vivanti AJ 2020 | Hospital | France | Pregnant woman admitted at 35+ weeks with COVID-19-like symptoms and subsequently tested & treated accordingly. | Case report |
| Von Kohorn I 2020 | Hospital | USA | Pregnant woman 34 weeks with vaginal bleeding & cramping, found to have thrombocytopenia, transaminitis, & hyperuricemia; infant delivered by C-section at 34 weeks (due to hemolysis, elevated liver enzymes, low platelet syndrome, suspicion for systemic COVID-19, & history of prior cesarean section) | Case report |
| Vouga M 2020 | International web registry | International | Women with symptoms for COVID-19 included in the study. Study conducted from 24 March to 26 July 2020. | Cohort study |
| Woodworth KR 2020 | Public health records | USA | 4,442 women with known pregnancy outcomes with laboratory-confirmed SARS-CoV-2 infection reported during 29 March to 14 October 2020. 4,495 live births. | Longitudinal surveillance |

| | | | | |
|---|---|---|---|---|
| Yang R 2020 | Maternal and Child Health Information System, Wuhan. | China | All pregnant women with singleton live birth recorded by the system between 13 January and 18 March 2020. | Cohort study. |
| Zaigham M 2020 | Hospital | Sweden | Case report of a pregnant woman admitted with COVID-19 symptoms & then diagnosis; subsequent investigations in the neonate. | Case report |
| Zhang L 2020 | Hospital | China | 18 patients with COVID-19 during late pregnancy, none critically ill. Epidemiological characteristics, clinical manifestations, laboratory tests, chest CT & pregnancy outcomes reported. | Case series (18 cases). |
| Zhang P 2020 | Case series of pathology of placental samples | USA | 364 consecutive women attending to give birth including 74 positive & 290 negative for SARS-CoV-2 by NP swab PCR. Placental pathology & clinical characteristics reported. | Case series (placenta analysis). |
| Zheng T 2020 | Hospital | China | Two pregnant women, admitted due to symptoms suggestive of COVID-19. (The only reported COVID-19 cases among pregnant women in this city during 20 January 2020, to 9 April 2020). Laboratory, imaging & SARS-CoV-2 nucleic acid tests were performed on the 2 women & their neonates | Case report (2 cases). |

Abbreviations: NP: nasopharyngeal

**Table 1b.** Results of included primary studies.

| Study ID | Patient numbers | Symptoms in mothers, including timing | Mode of delivery of babies | Sample sources: mother | Sample sources: neonate/ infant | Potential vertical tr, samples PCR-positive n/d for SARS-CoV-2 RNA unless otherwise stated | Live culture positivity | Other results |
|---|---|---|---|---|---|---|---|---|
| Alamar I 2020 | 1 mother & neonate dyad | Mother at 35+6 gestational age presented with vaginal bleeding & contractions; subjective fever, mild chills, fatigue, dysgeusia, anosmia beginning 1 day before presentation. | Urgent C-section due to bleeding | NP swab | NP swab; placental tissue biopsy | 1/1 | not attempted | Neonate positive at 24 hrs, 48 hrs & DOL 7. Mother & neonate remained asymptomatic during the 14 day follow-up period. Placenta ISH for SARS-CoV-2 RNA revealed strong signal in the villous syncytiotrophoblast; but no signal in villous stromal cells, Hofbauer cells, or villous endothelium |

| | | | | | | | | |
|---|---|---|---|---|---|---|---|---|
| Anand P 2020 | 69 mothers (39 tested positive) | Among 7 positive testing women: 1. critically ill & died 3 days postpartum, symptoms & timing NR; 2. fever 13 days pre-delivery, negative test day prior to delivery, time to diagnosis NR; 3. symptoms days 4 postpartum, time to diagnosis NR; 4. fever before delivery led to testing positive, time to diagnosis NR; 5. tested day 4 postpartum, symptoms NR; 6. symptoms day 12 postpartum; 7. day 10 symptoms so tested, time to diagnosis NR. | Elective C-section: 9/69; emergency C-section: 17/69. Remainder vaginal. | NP swab | NP/OP swab within 24h | 7/65; no deaths | not attempted | 2 maternal deaths attributed to COVID-19. All COVID-19 positive neonates remained asymptomatic to final follow-up at day 29. |
| Ayed A 2020 | 185 positive testing mothers | 88% of the patients had mild symptoms. Fever 58%, cough 51%. Time to diagnosis NR; tested on admission to maternity services. | 17/41: emergency C-section; remainder vaginal | NP swab | NP swab | 0/41 tested positive within 24h; 2/41 neonates tested positive on day 5; no deaths | not attempted | Cases assessed cases between 15 March and 31 May 2020; follow-up to 15 June 2020. At follow-up 98.8% of women had been discharged, median hospital stay 15 days. |

| | | | | | | | | |
|---|---|---|---|---|---|---|---|---|
| Bachani S 2020 | 348 pregnant women and 56 neonates | Most women that tested positive experienced mild infection (45/57; 78.9%), with one or two spikes of low-grade fever, cough, and/or diarrhoea that resolved in 2−3 days. Three (5.2%) women had moderate symptoms (fever and breathlessness). Timing of symptoms in relation to testing and/or delivery NR. | Spontaneous: 42; Induced labour: 15; Elective C-section: 9; Emergency C-section: 17 | NP swab | NP swab | | not attempted | 57/348 pregnant women positive; 3 deaths (comorbidities, 2 classed as COVID-19-related complications; 1 classed as COVID-19-related). 5/56 neonates positive; 2 tested positive within 24h; 2 tested positive on day 4; 1 tested positive on day 14 no deaths |
| Bandyopadhyay T 2020 | 1 mother & neonate dyad | Mild grade fever, pharyngeal swab taken that day and found positive for SARS-CoV-2 by RT-PCR; admitted to hospital for isolation then became asymptomatic one day later. Delivery 1 month after symptoms. Negative by PCR before delivery. | Vaginal | NP swab | NP swab | 1/1 | not attempted | Qualitative IgG serum antibody test positive; qualitative IgM serum antibody test negative On day 2 & day 3 of life, neonate tested negative. Mother asymptomatic after admission; tested negative 2 days before birth. |
| Barbero P 2020 | 91 women | Symptoms preceded & led to testing. Timing in relation to delivery NR. 40/91 developed pneumonia, and 4/91 required ICU admission | C-section: 11/23; remainder vaginal | NP swab or chest X-ray indicated | NP swab | 0/23 babies of women with active infection | not attempted | No suggestive symptoms in neonates on clinical follow-up; 1 neonate tested positive on day 8. |

| | | | | | | | | |
|---|---|---|---|---|---|---|---|---|
| Birindwa EK 2020 | 1 mother & neonate dyad | 3 weeks before admission, woman complained of fever, not responding to acetaminophen; fever persisted; swab test done 2 weeks later confirming SARS-CoV-2 positive by RT-PCR. She was admitted. 4 days later a rapid antigen test was negative for SARS-CoV-2. Preterm labour commenced and C-section was performed. | C-section | NP swab | NP swab | 1/1, neonate tested positive at birth, on day 3; died on day 5 due to sepsis | not attempted | Inflamed placenta |
| Bordbar A 2020 | 1 mother & neonate dyad | No fever, cough, dyspnea or GI symptoms | C-section | NP swab | NP swab; day 2, chest X-ray | 1/1; RT-PCR test on day 2, no symptoms; no symptoms at follow-up on day 14 | not attempted | |
| Chaudhary S 2020 | 26 pregnant women; 14 deliveries during study period | 16/26 positive tested women were symptomatic: 14/16 fever, 11/16 cough; 1 severe pneumonia leading to death. Timing of symptoms NR. | 3/14 vaginal, 11/14 C-section | NP swab | NP swab | 0/14 | not attempted | |

| | | | | | | | | |
|---|---|---|---|---|---|---|---|---|
| Cojocaru L 2020 | 31 neonates | 5/31 admitted to ICU. Other symptoms, and timing of symptoms, NR. | 8/31 C-section; 23/31 vaginal | Unclear, probably NP swab. | OP and NP swab at 24h and 48h | 0/31 | not attempted | Study on transmission and maternal bonding defined as rooming-in, skin to skin contact & breastfeeding. None of the neonates who bonded with their mothers tested positive. |
| Demirjian A 2020 | 1 mother & neonate dyad | Fever and respiratory symptoms on admission; condition deteriorated to severe, leading to C-section 7 days after admission. | C-section | Respiratory samples (day 6) and sputum (day 8) | NP swab, at birth and day 3, stool and blood samples day 3 | 1/1; RT-PCR negative at birth, RT-PCR respiratory sample positive on day 3, stool and blood samples negative | not attempted | Neonate no symptoms up to day 5, then developed fever (38.0°C), coryza, and mild tachypnea lasting 2 days. No respiratory support required. No symptoms at discharge on day 18. Mother on ventilation for 17 days. Discharged at 31 days. Genomic viral sequencing of SARS-CoV-2 virus isolated from samples from the mother's & the neonate's respiratory tract secretions. Genetic sequences were identical across the entire genome apart from a single-nucleotide difference between the baby and mother's genomes. |
| Farsi Z 2020 | 1 mother & 3 neonates | Woman had hypothyroidism, gestational hypertension (HTN), and gestational diabetes mellitus; also recent symptoms of cough, fever and myalgia 3 days before delivery. | Emergency C-section | PCR, test type and sample not specified | PCR, test type and sample not specified | 1/3; baby1: negative (day 3), ; baby2: negative day 3, PCR positive day 6, 23, PCR negative day | Not attempted | Neonate1: died from respiratory and gastrointestinal bleeding; Neonate2: healthy at follow-up; Neonate3: died day 16 of abdominal distension and enterocolitis |

| | | | | | | | | |
|---|---|---|---|---|---|---|---|---|
| | | | | | | 30, healthy at 37 day follow-up; baby3: PCR negative day 3,6 | | |
| Fenizia B 2020 | 31 mother & neonate dyads | 4/31 women had severe COVID-19 (defined by the need of urgent delivery for the deterioration of maternal conditions or by ICU/sub-intensive care admission). Radiological confirmation of interstitial pneumonia was obtained on admission or antepartum for all 4 severe cases and in 10 (32%) of the mild cases. | 25/31 vaginal; 6/31 caesarean | NP swab, placenta & umbilical cord biopsy, umbilical cord blood; amniotic fluid. | NP swab | 2/31, all babies healthy | not attempted | 2/31 maternal plasma samples positive (both had severe clinical outcome). SARS-CoV-2 in vaginal swab, placental tissue and cord plasma from 1 mother (severe outcome). SARS-CoV-2 in placental tissue from subject 1 mother. SARS-CoV-2 in one milk specimen only (severe clinical outcome). None of the tested 6 amniotic fluids, nor the 12 umbilical cords, resulted positive. Placentae from SARS-CoV-2 infected patients displayed a generalized immune activation profile compared to the uninfected |
| Ferraiolo A 2020 | 1 mother & neonate dyad | Hospitalized in order to undergo C-section. No symptoms on admission of fever/ respiratory/ GI symptoms, myalgia, malaise, ageusia/ anosmia | elective caesarean | NP swab placental biopsy and placental swab | NP swab at 0h and 24h | 0/1, healthy, discharged at 21 days | not attempted | Placental swabs COVID-19 positive, some placental inflammation on biopsy |

| | | | | | | | | |
|---|---|---|---|---|---|---|---|---|
| Gale C 2020 | 66 SARS-CoV-2 positive neonates (<12 HOL) | maternal symptoms NR | not reported | not specified | NP swab? - not specified for all | 2/unclear | not attempted | 17 babies born to mothers with confirmed infection within 7 days before or after birth. Only 2 babies considered vertical transmission due to positive nasopharyngeal swab |
| Gao W 2020 | 1 mother & neonate dyad | no fever or cough; thoracic CT showed no abnormality | emergency C-section | NP swab, | NP swab | 0/1; day 4 PCR test negative | not attempted | positive serum IgM and IgG antibody (colloidal gold method) were weak positive and strong positive |
| Gao X 2020 | Unclear, possibly 14 women | 11/14 women had fever, 6/14 had cough; all 14 had abnormalities evident by thoracic CT. 8/14 were given oxygen via nasal cannula; none was treated with respirator or mechanical ventilation. | 2 vaginal, 12 Caesarean section | OP or NP swab | nasal or oropharyngeal swab | 0/14 | not attempted | All breast milk samples negative for viral RNA, 3 breast milk samples positive for SARS-CoV-2 antibodies |
| Grimminck K 2020 | 1 mother & neonate dyad | Cough developed prior to admission | vaginal, induced labour | OP swab vaginal sample, urinary catheter sample, placenta | OP swab | 0/1 | not attempted | None of the samples taken from vagina, placenta, catheter or baby were positive |

| | | | | | | | | |
|---|---|---|---|---|---|---|---|---|
| He Z 2020 | 22 neonates | NR | not reported | not reported | Throat swab, amniotic fluid, umbilical cord blood, faeces, urine, blood. | 0/22 | not attempted | None of the samples taken from neonates tested positive for SARS-CoV-2. 3 newborns had elevated IgM Ab in umbilical cord & fetal blood; 12/22 had IgG positive for SARS-CoV-2; no deaths; 17/22 had hyperbilirubinaemia; report potential kidney damage in newborns |
| Hinojosa-Velasco A 2020 | 1 mother & neonate dyad | Fever, coughing, odynophagia, sore throat, headache, diarrhoea, rhinorhea, 2 days before admission | emergency C-section | NP and OP swab, breast milk and stool sample | nasopharyngeal and oropharyngeal swabs, during delivery, RT-PCR; stool sample | 1/1 | not attempted | Neonate considered to have a severe case of COVID-19 based on severity of symptoms, placed on oxygen. Day 4 milk & stool samples positive for SARS-CoV-2 RNA. 13 days after delivery, the infant's NP & OP swabs and stool samples were negative; maternal samples remained positive. |
| Hsu AL 2020 | 1 mother & neonate dyad | Myalgia 2 days prior to hospitalisation, no fever throughout hospitalisation | vaginal | placenta, RT-PCR and histology with immunohistochemical staining | not stated; RT-PCR | 0/1, SARS-CoV-2 RT-PCR test negative at 24h for baby (sample source not stated) | not attempted | IHC using SARS-CoV-2 nucleocapsid-specific monoclonal antibody demonstrated SARS-CoV-2 antigens throughout the placenta, under the umbilical cord, and at the central and peripheral placenta disc in chorionic villi endothelial cells, and rarely in CK7-expressing trophoblasts. |

| | | | | | | | | |
|---|---|---|---|---|---|---|---|---|
| Hu X 2020 | 6 mothers & 6 preterm neonates | In the 6 mothers, COVID-19 experienced 9, 10, 20, 13, 2, 2 days before delivery. All mild except one termed uncomplicated. Fever in 3/6, cough in 2/6. | 5 C-section; 1 vaginal | Unclear | Throat & anal swabs, blood; gastric aspiration before feeding, urine and stool samples. | 0/6 | not attempted | All samples of amniotic fluid, cord blood and breast milk negative. All neonate samples negative. |
| Khan MA 2020 | 66 mothers; 67 infants (2 twins) | Most of the 66 SARS-CoV-2-positive women were asymptomatic. One woman was admitted with respiratory failure. | 20 spontaneous vaginal delivery; 7 elective C-section; 40 emergency C- section | Not reported | NP swab | 0/67 | not attempted | |
| Kulkarni R 2020 | 1 mother & neonate dyad | Fever and myalgia one day prior to admission in active labour. Mother tested negative on admission and on day 5, serology negative on day 2, but serology positive on day 10. | Vaginal delivery, ventouse | NP swab | NP aspirate, cord stump, and placenta at birth | 1/1; | | All neonate samples positive at 12h.<br>Neonate admitted with fever, suspected sepsis. Negative on day 14, discharged on day 21. |

| | | | | | | | | |
|---|---|---|---|---|---|---|---|---|
| Liu W 2020 | 48 neonates | Fever (10/15, 67%) and cough (6/15, 40%) among mothers with COVID-19 diagnosis. | 42 C-section; 6 vaginal (no differences between the 3 groups of women) | NP swab, CT scan | amniotic fluid, placental swab, gastric lavage fluid at birth; neonatal serum,throat swab, and feces | 0/48. | | 15 neonates from COVID-positive women; 17 neonates from suspected COVID women; 16 neonates as control group. All samples from amniotic fluid, placental swab, gastric lavage fluid at birth; neonatal serum, throat swab, and feces negative. |
| Luo Q 2020 | 23 mother & neonate dyads | Fever (71%) and cough, 8/14 confirmed COVID-19 cases asymptomatic | C-section: 17; Vaginal: 6 | Throat swabs, breast milk, blood. | Throat swabs at delivery, serum at 1 month | 0/23 | not attempted | All throat swabs negative, all antibody tests at 1 month negative (8 infants). None of the women required ventilation. All breast milk samples negative for viral RNA; IgM antibody present in milk in 4 confirmed cases, IgG antibody negative in all samples |
| Lv Y 2020 | 1 mother & neonate dyad | Recurrent fever and cough over 2 weeks prior to admission | C- section | not stated, PCR test | Amniotic fluid, umbilical cord blood, placenta, and neonatal gastric fluid, pharyngeal and anal swab | 0/1 | not attempted | All neonatal samples negative. |

| | | | | | | | | |
|---|---|---|---|---|---|---|---|---|
| Maraschini A 2020 | 146 women, 143 singleton babies, 3 twins; 2 stillbirths, 147 live births | 41/146 (28.1%) of the COVID-19-positive women asymptomatic. 70/146 had fever, 68 had cough, 52 fatigue. Onset of clinical symptoms occurred in 9.5% of the cases on the day of delivery, and in 90.5% before it, the median value being 8 days (range 1-52 days). | Vaginal: 98; Elective C-section: 12; Emergency C-section (maternal or foetal indication): 25; Emergency C-section (COVID-19): 11 | NP swab | not stated, RT-PCR | 9/149 | not attempted | 9 infants tested positive for SARS-CoV-2, testing either at birth or up to 9 days after. Of 5 newborns with positive test within 24h, 4 were delivered vaginally, 1 by pre-labor C-section; none developed serious illness. 11 women received ventilation, no ECMO, no deaths. Two stillbirths at 30 and 35 weeks pregnancy. 23 infants admitted to NICU, 18 premature, no deaths |
| Marin Gabriel MA 2020 | 242 mothers, 248 neonates | Cough (33%); fever (30%). Odynophagia (2%) and chest pain (<2%%) were uncommon. 7 women required ICU. One mother died due to a massive thromboembolic event. | Vaginal: 179; Caeserean: 63 | NP and/or OP swab, blood | NP and/or OP swab | 11/230 positive (test at 18h). | not attempted | 2 additional cases at second test (48h)<br>222/248 neonates did not need respiratory support, no deaths. No cases of pneumonia or pneumonthorax. No additional cases at 1 month follow-up, 40% breast-fed exclusively at follow-up. |
| Masmejan S 2020 | 13 mother & neonate dyads | 5 asymptomatic, 6 mild, 1 severe and 1 critical. | Vaginal: 9; Forceps: 2; C-section: 2 | NP swab, placental swab. | NP swab, cord blood | 0/13; all samples (cord blood, placenta, neonate nasopharyngeal swab) negative | not attempted | 1 critical case amongst the women, requiring ventilation. No deaths; placental samples negative. |

| | | | | | | | | |
|---|---|---|---|---|---|---|---|---|
| Mohakud NK 2020 | 1 mother & neonate dyad | Presented with four-day history of low-grade fever, malaise, and breathing difficulty. | C-section | Not reported | Tracheal aspirate swab at 12 hours of life | 1/1 | Not attempted. | Woman developed breathing difficulty, decreased fetal movements, edema, and visual disturbance and was admitted to hospital & diagnosed with HELLP syndrome with hypothyroidism and moderate COVID-19 pneumonia. Neonate chest X-ray normal; neonate required ventilation at birth, subsequently experienced a seizure but recovered & was discharged healthy. |
| Moreno SC 2020 | 19 women, 21 neonates (including two sets of dichorionic diamniotic twins) | Women included in the study if symptomatic and with positive PCR for SARS-CoV-2. Cough 19/19; fever 7/19; shortness of breath 5/19. | 12 spontaneous vaginal; 7 C-section | NP swab | NP swab within first 24 hours of life. | 0/21 | Not attempted. | All 19 women presented with cough. Abnormal radiologic chest X-ray findings in 13/19 women. 8/19 births were premature deliveries. Among the neonates, no invasive mechanical ventilation was required. No neonatal sepsis or neonatal mortality was observed. |
| Nayak AH 2020 | 977 pregnant women: 141 with positive SARS-CoV-2 test; 836 with negative test.131 | 97% of the women were asymptomatic or had mild symptoms like fever or cough not requiring any oxygen therapy. (unclear if 97% is of all women in the study, or of all positive-testing women). | 481 vaginal (66/141 SARS-CoV-2 positive mothers); 443 C-section (67/141 SARS-CoV-2 positive | Unclear, probably NP swab. | Swab within first 24 hours of life | 3/131 | Not attempted. | Of 131 neonates tested, 3 tested positive on first swab within 24 hours of birth; all tested negative on day 5. |

| | | | | | | | | |
|---|---|---|---|---|---|---|---|---|
| | neonates tested. | | mothers); 10 instrumental (1/141 SARS-CoV-2 positive mothers) | | | | | |
| Ogamba I 2020 | 40 women, 25 deliveries within the time frame of the study; 23 neonates | 30/40 SARS-CoV-2-positive women reported <= one symptom due to COVID-19; 10/40 were asymptomatic. Symptoms included loss of smell & taste, nausea, vomiting, body aches, chills, headache, fever, dry cough, shortness of breath, abdominal pain, difficulty breathing, chest pain. | 17/25 vaginal; 8/25 C-section | NP or nasal swab | NPl or nasal swab | 0/20 | Not attempted. | |
| Olivini N 2020 | 5 mother & neonate dyads | 3/5 women reported symptoms: 1 low grade fever, 2 anosmia, 1 dysgeusia, 1 musculoskeletal pain. Onset of symptoms occurred at day 1, 5 and 19 after childbirth. | 2 vaginal, 3 elective C-section | rhino-pharyngeal swabs | rhino-pharyngeal and rectal swabs | 0/1 neonate tested within 24h; 5/5 tested positive, at day 2, 16, 12, 6 and 4 | Not attempted. | Neonates were asymptomatic or paucisymptomatic, no fever or respiratory symptoms. Three women reported symptoms: 1 low grade fever, 2 anosmia, 1 dysgeusia, 1 musculoskeletal pain. Onset of symptoms occurred at day 1, 5 and 19 after childbirth. Single samples of breastmilk from each of two of the SARS-CoV-2 positive mothers tested negative by RT-PCR. |

| | | | | | | | | |
|---|---|---|---|---|---|---|---|---|
| Oncel MY 2020 | 125 pregnant women; NP swabs available for 120 newborns (not possible for 5 [asymptomatic] newborns) | 85/125 SARS-CoV-2 positive women had at least one COVID-19 symptom. (40/125 had had close contact.) 8 were admitted to ICU for mechanical ventilation, 6/these 8 died. | 89 C-section, 36 other (mode not reported) | Not reported | NP/pharyngeal; subsequently, deep tracheal aspirates | 4/120; no NP sample tested positive within 24h; 1 positive on day 2, 2 positive on day 4; 1 deep tracheal aspirate tested positive on day 1 | Not attempted. | Additional COVID-19 tests done in some (rationale not recorded): placenta tissue (n: 5), amnion fluid (n: 4), deep tracheal aspirate (n: 9), serum (n: 3), stool (n: 2), and breast milk (n: 6), which were all negative for SARS-CoV-2 except only one positive deep tracheal aspirate. 8/125 mothers were admitted to ICU for mechanical ventilation; 6 died. |
| Palalioglu RM 2020 | 1 mother and neonate dyad | asymptomatic | C-section | NP swab | NP swab | 0/1 | Not attempted. | CT performed after C-section showed ground glass opacities in both lungs of the mother despite no COVID-19 symptoms. Baby was admitted to the ICU after developing feeding intolerance and vomiting 6 h after being breastfed, later discharged healthy. Also amniotic fluid, cord blood and placenta, postoperative breast milk were negative by RT-PCR. |

| | | | | | | | | |
|---|---|---|---|---|---|---|---|---|
| Parsa Y 2020 | 1 mother & neonate dyad | Presented with signs and symptoms of acute respiratory illness including shortness of breath and cough. | C-section | NP swab | Not reported | 1/1 24 hours after delivery | Not attempted. | Mother aged 41 years, with opium addiction, with signs and symptoms of acute respiratory illness presenting with labor pain & vaginal leak at 37 weeks of gestation. CT-scan revealed ground-glass opacities. The RT-PCR results of the amniotic fluid and neonate (less than 24 hours after birth) were positive for COVID-19. The newborn suffered vomiting during the first 24 hours after birth, due to opium withdrawal syndrome. She was admitted to the NICU and received supportive care without need for respiratory support. |
| Pereira A 2020 | 22 mother & neonate dyads | 6/22 mild symptoms, 5/22 pneumonia, 10/22 no symptoms | 4 C-section, 18 vaginal (4 of which instrumental) | Not reported. | NP swab taken within first two hours of life. | 0/22 | Not attempted. | Two born preterm needed admission to the NICU. During follow-up period, there were no major complications, and no neonates were infected during breastfeeding. |
| Pessoa FS 2020 | 1 mother & neonate dyad | Admitted with flu-like symptoms including fever, dry cough & fatigue; symptom onset 7 days prior to admission, progressed to dyspnoea | C-section | Not reported | Swab at 6 HOL, source not reported. | 1/1 | Not attempted. | |

| | | | | | | | | |
|---|---|---|---|---|---|---|---|---|
| Pissarra S 2020 | 10 mother & neonate dyads | 7/10 symptomatic; 1 mother admitted to ICU; days from beginning of symptoms to delivery ranged from 0-15. No details of symptoms or severity | 6 vaginal, 4 C-section (obstetrical indication) | NP swab or orophary ngeal swab | NPI swab and bronchial secretions | 0/10 | Not attempted. | All neonates separated from mothers at birth and transferred to NICU. All remained symptom free and tested negative for SARS CoV2 at birth & at 48 h of life. |
| Popofsky S 2020 | 160 neonates | 59 mothers (36.9%) were symptomatic during perinatal hospitalization, with fever (>37.7°C), cough, shortness of breath, or a combination of these. 25 mothers (15.6%) had been symptomatic before the perinatal admission but were no longer symptomatic during hospitalization. 76 (48%) were never symptomatic. Mean day of onset of symptoms 14. | 122 vaginal; 38 C-section | NP swab | NP swab | 1/ 160 newborns (positive test was on day of life 5, after a negative test at birth) | Not attempted. | Mother of infant testing positive was symptomatic at the time of birth & had close contact with the newborn whilst symptomatic. |
| Pulinx B 2020 | 1 mother & two twin neonates | Presented to ED at 22w with rhinitis and fever (39.2 °C) | Vaginal | NP swab | placental and amniotic fluid | 2/2 (placental and amniotic fluid) | Not attempted. | Both fetuses died prepartum. Placental histological examinations showed chronic intervillositis and extensive intervillous fibrin depositions with ischemic necrosis of the surrounding villi. Viral localization in the placental syncytiotrophoblast cells was confirmed by immuno-histochemistry. No other cause of fetal demise was identified. |

| | | | | | | | | |
|---|---|---|---|---|---|---|---|---|
| Rebello CM 2020 | 1 mother & neonate dyad | Anosmia, ageusia, runny nose, dry cough, headache & myalgia, no fever (10 days before delivery); diagnosed with COVID-19 eight days before delivery | Vaginal | NP swab | umbilical cord blood and skin swabs | 1/1 (cord blood and neonate skin swabs) | Not attempted. | |
| Rivera-Hernandez P 2020 | 1 mother & neonate dyad | Presented at 33weeks pregnant, 3-day history of malaise & dyspnoea; ICU admission & ventilation; delivery on day 4 of admission | C-section | NP swab | NPI swab | 1/1 | Not attempted. | No postdelivery contact of neonate with mother or father; father tested negative. Chest radiography in neonate showed mild ground glass appearance & increased pulmonary vascularity bilaterally, consistent with respiratory distress syndrome. Neonate treated in NICU. |
| Rubio Lorente AM 2020 | 2 pregnant women | Case 1, symptomatic; Case 2, fever 2 weeks before, resolved prior to amniocentesis | n/a | serology & NP swab; amniocentesis | amniotic fluid | 0/2 | not attempted | Amniotic fluid negative for SARS-CoV-2 by RT-PCR for both cases |
| Sajjan GR 2020 | 1 mother & neonate dyad | Asymptomatic of respiratory disease | C-section | not reported | not reported | 1/1, IgG | Not attempted | Neonate Covid Ig G positive, Neonatal lower limb gangrene considered to result from in utero transmission of SARS-CoV-2 infection. At 15 days, neonate was stable. |

| | | | | | | | | |
|---|---|---|---|---|---|---|---|---|
| Schwartz DA 2020 | 19 neonates who tested positive and their mothers. | Predelivery symptoms: 10 respiratory symptoms (2 severe), 4 asymptomatic, 5 no information reported. | 11 births by C-section; 5 vaginal; 3 mode of delivery not reported | Not reported | NP, endotracheal or OP swab | | Not attempted | 5/19 mothers negative despite positive neonate. 3 infants readmitted later due to signs of possible COVID-19. One of the included 19 neonates was one of triplets, the other two neonates had died of respiratory distress. |
| Shende P 2020 | 1 mother & first trimester fetus | Asymptomatic, tested positive 6 weeks previously. | Termination at 13 weeks | Throat swab | Products of conception, serum, placenta, amniotic fluid | 1t | Not attempted | Products of conception negative for IgM. Placenta showed infection.Viremia persisted in the placenta six weeks after the mother was tested positive. |
| Singh MV 2020 | 1 mother & neonate dyad | Asymptomatic | C-section | NP swab | NP swab | 1/1 | Not attempted | C-section performed at 38 weeks due to positive SARS-CoV-2 test. Neonate healthy. NP swab turned negative on day 10 of life. |
| Sisman J 2020 | 1 mother and infant dyad | 3 days pre-delivery admitted with fever and diarrhoea (admitted due to possible preterm labour) | vaginal | NP swab | NP swab | 1/1 at 24 and 48 HOL | | Premature rupture of membranes so labour induced. Hyperbilirubinemia in the neonate. Neonatal fever & respiratory distress at day 2, resolved by day 5. Chest radiograph normal. Discharged as healthy at day 21. |

| | | | | | | | | |
|---|---|---|---|---|---|---|---|---|
| Smithgall MC 2020 | 76 pregnant women | 26/51 SARS-CoV-2-positive women were asymptomatic; 32 had cough, 27 had fever, 14 had myalgia, 6 had sore throat, 6 had fatigue. 4 suffered severe disease. | 51/76 vaginal; 25/76 C-section | placentas | NR | 0/51 | Not attempted | No adverse perinatal outcomes; no specific histomorphologic changes in placentas; no evidence of direct viral involvement identified by ISH and IHC. Evidence of maternal–fetal vascular malperfusion was identified: SARS-CoV-2 positive women's placentas significantly more likely to show villous agglutination & subchorionic thrombi than SARS-CoV-2-negative women's placentas. |
| Stonoga E 2020 | 1 mother & fetus dyad | Dyspnea, dry cough, high temperature (38.5°C), anosmia, nausea, vomiting, and diarrhea had developed 2 days before hospitalization. | C-section | NP swab tested by RT-PCR | Amniotic fluid (before amniotic membranes ruptured), umbilical cord blood, placental membranes, & cotyledon fragments | 0/1 | Not attempted | Fetal death. All fetal samples tested negative. Multifocal chronic histiocytic intervillositis in the placenta, potentially relevant to cause of death of the fetus, but other causes not excluded. |

| | | | | | | | | |
|---|---|---|---|---|---|---|---|---|
| Tang F 2020 | 20 mother & neonate dyads | Unclear. However, the mothers of two neonates who tested positive for RT-PCR at 1–2 days after birth also tested positive & exhibited typical symptoms such as fever, cough, one day pre delivery | 5 vaginal, 15 C-section | Unclear | Throat swabs, serum, amniotic fluid, umbilical blood (and radiology) | 3/20 | Not attempted | No neonate had a fever. Radiological examination showed that 10 neonates, eight neonates, and two neonates were diagnosed with pneumonia, increased lung markings, and no abnormality in the lungs, respectively. All amniotic fluid & umbilical cord blood tested negative. |
| Tang J 2020 | 2 mother & neonate dyads | Case 1: fever & respiratory symptoms 13 weeks prior to delivery, symptoms resolved; admission to hospital 13 weeks later with premature rupture of membranes, no respiratory symptoms, but serum antibodies positive for SARS-CoV-2. Case 2: mild respiratory symptoms predelivery. | 1 vaginal birth, 1 birth by C-section | Throat swabs, serum | Throat swabs at birth, day 3, day 7. Serum on day 7. | 0/2 by PCR; 2/2 positive by IgG, 1/2 positive by IgM. | Not attempted | Mothers: throat swabs negative by PCR, 2/2 were positive for serum IgM, 1/2 positive for IgG antibodies. The two mothers did not develop serious complications; outcomes in the neonates were good. Neonate serum IgM antibody to SARS-COV-2 was negative and IgG was positive on the 7th day after birth. |
| Vendola N 2020 | 2 mother & neonate dyads | Both cases asymptomatic | 1 vaginal, 1 C-section | NP swab, serum | NP swab, serum, umbilical cord blood, peripheral blood soon after delivery. | 0/2 for SARS-CoV-2 RNA in throat swabs; 2/2 for IgG in neonatal serum | Not attempted | |

| | | | | | | | | |
|---|---|---|---|---|---|---|---|---|
| Vinuela MC 2020 | 100 asymptomatic pregnant woman & their neonates | All asymptomatic | 94 vaginal, 6 C-section | NP swab | Unclear | 0/100 | Not attempted | No fetal transmission was observed (details NR). Maternal & neonatal prognosis excellent. All Cts from maternal swabs were >35. |
| Vivanti AJ 2020 | 1 mother & neonate dyad | Fever (38.6 °C) and severe cough | C-section | Blood, NP swab, vaginal swab, amniotic fluid during C-section. | NP and rectal swabs at 1 hour, 3 and 18 days. Cerebrospinal fluid sample at 3 days. | 1/1 | Not attempted | Mother admitted at 35+2 weeks of gestation with fever (38.6 °C), severe cough, abundant expectoration for 2 days before hospitalisation. Neonatal blood culture was negative for bacteria or fungi. All neonatal swab samples tested positive by PCR. CSF sample negative. Placenta histology some positive Ab staining for SARS-CoV-2 |
| Von Kohorn I 2020 | 1 mother & neonate dyad | Maternal gestational diet-controlled diabetes. Presented with cough, vaginal bleeding at 34 weeks gestation. COVID-19 positive 14h prior to delivery. Mother had a cough for 1 week prior to NP swab & test. | C-section | NP swab | Cord blood at birth. NP swabs at 24 hours and 49 HOL. Neonatal blood & urine over first 7 days of life. | 1/1 (several consecutive samples tested, some were positive by PCR with variable Ct values) | Not attempted | Infant asymptomatic with normal laboratory studies but some NP swabs tested positive for SARS-CoV-2 RNA. Placenta histology showed no direct evidence of virus. |

| | | | | | | | | |
|---|---|---|---|---|---|---|---|---|
| Vouga M 2020 | 1,033 pregnant women: 926 tested positive, 107 tested negative. 384 neonates tested at birth. | Symptoms included fever, anosmia, cough, sore throat, dyspnoea, myalgia, fatigue, headache, nausea. Symptom frequency reported in the cohort, symptom severity not reported. Some asymptomatic | Among 926 SARS-CoV-2 positive women: 469 vaginal, 256 C-section, 6 unknown; among 107 SARS-CoV-2 negative women: 44 vaginal; 22 C-section, 1 unknown. | NP swab | NR | 11/384 | Not attempted | No difference in obstetrical and neonatal outcomes were observed between positive- and negative-testing women. |
| Woodworth KR 2020 | 4,442 positive pregnant mothers, 4,495 live births. | Symptom status was known for 2,691 (60.6%) women (including women with symptoms reported on COVID-19 form), 376 (14.0%) of whom were reported to be asymptomatic. Symptoms and severity not reported | 2,589 vaginal, 1,331 C-section. Among 3,920 women testing positive for SARS-CoV-2: 2,589 vaginal, 1,331 C-section. | NR | NR | 16/610 | Not attempted | % positivity was 4.3% (14 of 328) among infants born to women with documented infection ≤14 days before delivery and 0% (0 of 84) among those born to women with documented infection >14 days before delivery. Molecular testing results available for 610 infants. Other perinatal outcomes also reported. |

| | | | | | | | | |
|---|---|---|---|---|---|---|---|---|
| Yang R 2020 | 11,078 live singleton birth mother & neonate dyads: | Those with fever and cough or having abnormal CT scan were tested for SARS-CoV-2. Severity of symptoms not reported. | 5006/11,078 vaginal, 6,072 C-section (Among 65 SARS-CoV-2 positive women, 13 vaginal, 52 C-section) | Pharyngeal swab | Unclear, probably pharyngeal swab as for mother. | 0/38 | Not attempted | 65/11,078 mothers tested positive during pregnancy. 38 /58 newborns of positive testing mothers were tested for SARS-CoV-2 after birth: 0 positive. OR for perinatal conditions & SARS-CoV-2 positivity also reported. |
| Zaigham M 2020 | 1 mother & neonate dyad | 34 weeks pregnant. Fever, dry cough, abdominal pain. Treated in postpartum ward (no ICU) | C-section | NP & throat swab | NP swab 48 hours after delivery | 1/1 | Not attempted | High viral load in placenta with massive perivillous fibrin deposition, acute intervillositis in areas with strong positivity for SARS-CoV-2 & chorangiosis in the areas less affected by infection and inflammation. Whole genome sequencing of isolates from the mother & placenta revealed a single variant of the virus. |
| Zhang L 2020 | | 35 to 41 weeks pregnant. Most positive pregnant women had fever, cough, sore throat, fatigue, chest tightness or shortness of breath, diarrhea, and runny nose. Clinical typing: 1 case "mild", 16 cases "ordinary", 1 case "severe" | 1 vaginal, 17 C-section | Throat swab | Sample source NR | 0/18 | Not attempted | 5/18 neonates diagnosed with bacterial pneumonia, successfully treated. |

| | | | | | | | | |
|---|---|---|---|---|---|---|---|---|
| Zhang P 2020 | 364 pregnant women | Not reported | 137/364 vaginal, 227/364 C-section; among SARS-CoV-2 positive women 54/74 vaginal, 20/74 C-section. | NP swab | Placental tissue | 2/53 by in situ hybridization in placenta | Not attempted | 74/364 women tested positive. 2/53 placental samples positive from PCR positive mothers. 0/10 neonatal placentas positive from negative mothers. No histolopathological features within the placentas specific to maternal SARS-CoV-2 infection. |
| Zheng T 2020 | 2 x mother + neonate dyads | Ppt 1: 36weeks pregnant, fever & asthenia at admission, CT showed bilateral lung infection, classified as "severe COVID-19"; Ppt 2: 39weeks, admitted due to abnormal chest CT; no fever or cough, CT showed bilateral lung infection. | C-section (both) | Throat swab | Throat swab, anal swab, urine, &blood; sample timing unclear | 0/2 | Not attempted | |

**Table 1c.** Summary of findings from reviews

| Study | Fulfils systematic review methods (Y/N/U); Risk of bias tool used | Stated research question | Databases and search date | No. included studies (No. participants) | Main results | Authors' key conclusions |
|---|---|---|---|---|---|---|
| Akhtar H 2020 | Yes; RoB: Newcastle Ottawa Scale | Review of association of SARS-CoV-2 infections with pregnancy, foetal, and neonatal outcomes | PubMed, Scopus, Medline, Cochrane database, Google Scholar from 1 December 2019 to 22 May 2020 | 22 (156 pregnant women with COVID-19; 108 neonates) | Most common maternal/foetal complications included intrauterine/foetal distress (14%) and premature rupture of membranes (8%). Neonatal clinical manifestations of COVID-19: shortness of breath (6%), gastrointestinal symptoms (4%), fever (3%). | COVID-19 infection in pregnancy leads to increased risk in pregnancy complications such as preterm birth, PPROM; may rarely lead to maternal death. No evidence to support vertical transmission of SARS-CoV-2. Limited evidence on impact of COV)D-19 on newborns. |
| Al Qahtani MA 2020 | No | Summarize research regarding pregnancy and vertical transmission of COVID-19 | Pubmed from 1 January 2020 to March 20 2020 | 9 studies (74 pregnant women, 72 positive for COVID-19, 2 negative but symptomatic; 75 neonates) | All except one neonate tested negative; one tested positive for throat swab, but negative for cord blood and placenta. | Only one infant from a positive mother tested positive for COVID-19 throat swab. No difference between pregnant and nonpregnant patients regarding clinical manifestations. |
| Amaral W 2020 | Yes; RoB: GRADE | To investigate pregnant women infected with COVID-19 in terms of signs and symptoms, type of delivery, comorbidities, maternal and neonatal outcomes, and the possibility of vertical transmission. | Embase and PubMed search on 31 October 2020. | 70 studies (1457 pregnant women) | Maternal signs and symptoms: fever, cough, nausea. Maternal and fetal outcomes: premature birth (n = 64), maternal death (n = 15), intrauterine fetal death or neonatal death (n = 16), cases of intrauterine fetal distress (n = 28), miscarriage (n = 7), decreased fetal movements (n = 19), severe neonatal asphyxia (n = 5). 39 newborns positive for SARS-CoV-2. SARS-CoV-2 RNA detected in placenta (n = 13) and breast milk (n = 6). | COVID-19 during pregnancy can result in maternal, fetal, and neonatal complications. SARS-CoV-2 viral exposure of neonates during pregnancy and delivery cannot be ruled out. Need for long-term follow-up. |

| | | | | | | |
|---|---|---|---|---|---|---|
| Bellos I 2020 | Yes; RoB: Quality appraisal by domains, tool not stated | Evaluate maternal and neonatal outcomes in COVID-19 pregnancies, factors associated with perinatal viral transmission. | Medline, Scopus, CENTRAL, Web of Science and Google Scholar databases up to 3 June 2020. | 60 studies | Maternal symptoms: Fever, cough, shortness of breath, 15% asymptomatic. Severe disease in 7-11%. 2 maternal deaths. Neonatal symptoms: fever, shortness of breath, vomiting. 20% asymptomatic. No difference in neonatal transmission rate between women with and without severe disease (OR: 1.94, 95 % CI: 0.50–7.60). Preterm birth in 16-30%. Stillbirth, 3 cases, neonatal deaths, 2 cases. Vertical transmission, 4 cases. | Maternal and neonatal clinical course typically mild, low mortality rates. Risk of vertical transmission low, not linked to maternal disease severity.. |
| Bwire GM 2020 | Unclear; RoB: No quality assessment, as majority studies case reports. | Determine likelihood of vertical transmission in COVID-19 exposed infants; whether exposed negative infants had SARS-CoV-2 antibodies | PubMed/MEDLINE, Google Scholar. Websites of WHO, CDC; Google search of specific journals; Research Square, medRxiv. From 1 December 2019 to 18 May 2020 | 33 studies (205 infants born to COVID-19 positive mothers) | 6.3% (13/205; 95% CI: 3.0%–9.7%) infants tested positive for COVID-19 virus at birth. SARS-CoV-2 IgG/IgM in 90% COVID-19-negative infants (10/11; 95% CI: 73.9%–107.9%) (6 studies). Median antibody levels IgG 75.49 AU/ml (range, 7.25–140.32 AU/ml), IgM 3.79 AU/ml (range, 0.16–45.83 AU/ml) | Low possibility of vertical transmission of COVID-19, SARS-CoV-2 antibodies in vertically exposed but negative infants. |
| Caparros-Gonzalez RA 2020 | No | Guide clinicians on management of pregnant women re maternal-fetal-neonatal SARS-CoV-2 infections and breastfeeding during COVID-19 pandemic | Web of Science, PubMed, Scopus, Dialnet, CUIDEN, Scielo, Virtual Health Library from 25 August 2020 to 10 November 2020. | 49 studies (329 pregnant women and 331 neonates) | Samples positive for SARS-CoV-2 RNA: 15 placental swabs positive on fetal side of the placenta; 7 breastmilk; 1 umbilical cord; 1 amniotic fluid | Evidence supports potential of congenital, intrapartum, and postnatal maternal-fetal-neonatal SARS-CoV-2 infections. Mothers should follow recommendations e.g. wearing facemask and hand washing before and after breastfeeding. |
| Cavalcante de Melo 2020 | Yes; RoB: Newcastle Ottawa Scale | Analyze whether COVID-19 in pregnant women is related to premature birth and birth weight; summarize diagnostic results of neonates born to mothers with COVID-19 to investigate possibility of vertical transmission. | PubMed, Scopus, LILACS, Web of Science, Google Scholar, Preprints, bioRxiv, and medRxiv. Search date 4 May 2020 | 8 studies (279 women, 60 diagnosed with COVID-19) | No significant association between COVID-19 and preterm delivery (OR = 2.25; 95%CI: 0.96, 5.31; p = 0.06; $I^2$ = 0%). No significant relationship between birth weight and COVID-19 (MD = -124.16; 95%CI: -260.54, 12.22; p = 0.07; $I^2$ = 0%). | COVID-19 in pregnant women may not be associated with occurrence of preterm deliveries or birth weight of newborns. Level of evidence low. A few reports suggest vertical transmission possible, but evidence uncertain. |

| | | | | | | |
|---|---|---|---|---|---|---|
| Centeno-Tablante E 2020 | Yes; RoB: Newcastle Ottawa Scale. | Summarise information on breast milk and breastfeeding to inform guidance during COVID-19 pandemic. | MEDLINE (PubMed), the WHO COVID-19 database, Cochrane Library, Web of Science Core Collection, Embase. COVID-19 subset of WHO International Clinical Trials Registry Platform. Search date 7 July 2020 | 37 studies (77 breast-feeding mothers) | 19/77 children confirmed COVID-19 cases (positive RT-PCR). 9/68 breast milk samples from mothers with COVID-19 were positive for SARS-CoV-2 RNA; of the exposed infants, 4 were positive, 2 were negative for COVID-19. | No evidence of SARS-CoV-2 transmission through breast milk. Long-term follow-up studies needed |
| Chi J 2020 | Yes; RoB: Case report/case series (Murad et al.) | Summarise clinical features and maternal–infant outcomes of pregnant women infected with COVID-19 and their infants, including possibility of vertical transmission. | PubMed, Embase, Medline, MedRxiv, CNKI, and the Chinese Medical Journal Full Text Database. Search up to 18 April 2020. | 20 studies (230 women with COVID-19, 156 newborns) | Pregnant women: 34.6% obstetric complications, 59% fever, 40.7% lymphopenia, 5.19% received mechanical ventilation, 7 critically ill. Deaths: 1 maternal, 2 neonatal.<br>Newborns: 24.74% premature, 5 SARS-CoV-2 positive (throat swab), all positives delivered by C-section. 8 negative newborns with elevated SARS-CoV-2 IgM and IgG. PCR tests of vaginal secretions, breast milk, amniotic fluid, placental blood, placental tissues all negative. | Most pregnant patients mildly ill. Mortality of pregnant women with COVID-19 lower than for COVID-19 patients overall. C-section more common than vaginal delivery for pregnant women with COVID-19. Main newborn adverse event: premature delivery. Vertical transmission rate 3.91%. |
| Della Gatta AN 2020 | Yes; RoB: Case report/case series (Murad et al.) | Review clinical outcomes for pregnant patients with COVID-19. | PubMed, CINAHL, Scopus. From 14 March 2020 to 16 March 2020 | 6 studies (51 pregnant women) | 48 pregnant women, 46 cesarean (96%), 2 vaginal delivery. 1 stillbirth and 1 neonatal death. Median gestational age 36.5 weeks (interquartile range, 35–38), 15 preterm birth (39%); indications for cesarean delivery not clearly described. | Vertical transmission of SARS-CoV-2 excluded thus far, outcomes for mothers and neonates generally good; high rate of preterm delivery by cesarean, mostly elective, reason for concern. Reasonable to question whether cesarean delivery for pregnant patients with COVID-19 warranted. |
| Deniz M 2020 | No | Review evidence on vertical transmission of SARS CoV-2. | PubMed, Global Health (OVID), SCOPUS from 1 February 2020 to 1 June 2020 | 50 studies (714 pregnant women with COVID-19, 606 neonates) | 17 newborns testing positive for SARS CoV-2 by RT-PCR. 3 neonates with elevated SARS-CoV-2 IgG and IgM. Reporting samples testing positive: 8 placental tissues, 3 breast milk, 1 amniotic fluid | Possible vertical transmission of SARS CoV-2 has been observed in some studies. More RT-PCR tests on amniotic fluid, placenta, breast milk and cord blood are required. |

| | | | | | | |
|---|---|---|---|---|---|---|
| Dhir SK 2020 | Yes; RoB: Newcastle Ottawa Scale | Synthesize literature on various modes of transmission (congenital, intrapartum, and postpartum), clinical features and outcomes of SARS-CoV-2 infection in neonates. | PubMed, EMBASE, Web of Science until 9 June 2020. | 86 studies (1992 pregnant women, of which 1125 gave birth to 1141 neonates) | 281 (25%) neonates were preterm, preferred delivery mode caesarean section (66%). 41 case reports of 43 mother-baby dyads reporting 16 preterm births, 9 low birth weight. 58 neonates reported with SARS-CoV-2 infection, 29 (50%) symptomatic (23 required ICU). 70% with respiratory symptoms. No neonate mortality. | Limited low-quality evidence suggests risk of SARS-CoV-2 infections in neonates is extremely low. Most COVID-positive neonates symptomatic and required intensive care. Postpartum acquisition the commonest mode of infection in neonates, a few cases of congenital infection. |
| Di Toro F 2020 | Yes; RoB: Joanna Briggs Institute | Assess the impact of COVID-19 on maternal and neonatal outcomes. | PubMed, EMBASE, MedRxiv, Scholar, Scopus, Web of Science database up to 8th May 2020. | 24 studies (1100 pregnancies). | Pooled prevalence of pneumonia 89% (95%CI 70–100), prevalence of women admitted to ICU 8% (95%CI 1–20). 3 stillbirths, 5 maternal deaths. Pooled prevalence of 85% (95%CI 72–94) for caesarean deliveries. 3 neonatal deaths. Prevalence of COVID-19-related admission to neonatal ICU 2% (95%CI 0–6). 19/444 neonates positive for SARS-CoV-2 RNA at birth. 1 case of elevated levels of IgM and IgG, but negative swab. | Adverse outcomes e.g. ICU admission or patient death can occur, but clinical course of COVID-19 in most women not severe, infection does not significantly influence the pregnancy. High caesarean delivery rate reported, but no clinical evidence to support this. In most cases disease does not threaten the mother, vertical transmission not clearly demonstrated. COVID-19 should not be considered indication for elective caesarean section. |

| | | | | | | |
|---|---|---|---|---|---|---|
| Diriba K 2020 | Yes; RoB: Joanna Briggs Institute | Assess the effect of coronavirus infection (SARS-CoV-2, MERS-CoV, and SARS-CoV) during pregnancy and its possibility of vertical maternal–fetal transmission. | PubMed, Web of Science, Embase, Google Scholar, Cochrane Library until the end of April 2020 | 39 studies (1316 pregnant women) | Most common clinical features: fever, cough, myalgia (prevalence 30 to 97%); Abnormal lab findings: lymphocytopenia, CRP (55–100%). Pneumonia most diagnosed clinical symptom of COVID-19 and non-COVID-19 infection (prevalence 71-89%). CT imaging: Bilateral pneumonia (57.9%), ground-glass opacity (65.8%). Most common treatment: hydroxychloroquine (79.7%), ribavirin (65.2%), oxygen therapy (78.8%). Maternal outcome: rate of preterm birth < 37 weeks 14.3%, preeclampsia 5.9%, miscarriage 14.5%, premature rupture of membranes 9.2%, fetal growth restriction 2.8%. COVID-19 pregnant women: 56.9% delivered by cesarean, 31.3% admitted to ICU, 2.7% deaths. Perinatal outcomes: fetal distress 26.5%, neonatal asphyxia 1.4%, apgar score < 7 at 5 min 1.2%, neonates admitted to ICU 11.3%, death 2.2%. No reports of in utero transmission of CoV. | Coronavirus infection more likely to affect pregnant women. Respiratory infectious diseases demonstrated increased risk of adverse maternal obstetrical complications than general population. No studies reported transmission of CoV from the mother to the fetus in utero, may be due to very low expression of angiotensin-converting enzyme-2 in early maternal–fetal interface cells. |
| Dube R 2020 | No | Review literature to determine effects of confirmed cases of COVID-19 in pregnant women by estimation of mother to child transmission, perinatal outcome and possible teratogenicity. | PubMed, Embase, LitCovid, Google Scholar, EBSCO MEDLINE, CENTRAL, CINAHL, MedRXiv, BioRXiv, Scopus between 1 November 2019 and 10 August 2020 | Mother to child transmission: 72 studies (1408 neonates); Perinatal outcomes: 60 studies (1318 foetuses) | Positive tests for SARS-CoV-2 RNA: 3.67% of neonates (NP swab) cord blood 7.1%, placenta 11.7%, amniotic fluid 6.8%, faecal and rectal swabs 9.6%. Rate of preterm labour 26.4% (OR=1.45, 95% CI 1.03 to 2.03), caesarean delivery 59.9% (OR=1.54, 95% CI 1.17 to 2.03). Most common neonatal symptom breathing difficulty (1.79%). Stillbirth rate (COVID-19 positive mothers) 9.9 per 1000. Congenital transmission rate 9/1408 (0.63%). | Chances of mother to child transmission of the SARS-CoV-2 virus is low. Perinatal outcome for the foetus is favourable. Increased chances of caesarean but not preterm delivery. Stillbirth and neonatal death rates low. No reported congenital anomalies. |

| | | | | | | |
|---|---|---|---|---|---|---|
| El-Wahab EWA 2020 | No | Summarize SARS-CoV-2 transmission and provide scientific support for the prevention and control of COVID-19. | ISI Web of Knowledge, PubMed, Medline, ScienceDirect, EMBASE, EBSCO, ProQuest, Google Scholar, WHO, CDC, Mayo Clinic, bioRxiv, medRxiv, SSRN, Qeios from 28 December 2019 to 31 July 2020 | 302 studies | Broad review on various modes of transmission. Regarding vertical transmission, report generally on possibility of mother-to-child transmission and detection of SARS-Co-V-2 in breastmilk. | Very broad review of all types of transmission, including possible mother-to-child and breastmilk. |
| Figueiro-Filho EA 2020 | No | Identify the most significant studies reporting on COVID-19 during pregnancy and provide an overview of SARS-CoV-2 infection in pregnant women and perinatal outcomes. | PubMed, EMBASE, Cochrane Library, Google Scholar until July 20, 2020. | 8 studies (10,966 COVID-19 pregnant women) | Pregnant women not more affected by respiratory complications of COVID-19, compared to general population. Down-regulation of ACE2 receptors induced by SARS-CoV-2 cell entry might have been detrimental in subjects with pre-existing ACE2 deficiency associated with pregnancy, which may explain the worse perinatal outcomes. | Maternal characteristics, clinical symptoms, maternal and neonatal outcomes of 10,996 cases of COVID-19 and pregnancy are not worse or different from the general population. Pregnant women are not more affected by the respiratory complications of COVID-19, compared with non-pregnant available data. |
| Gao Y 2020 | Yes; RoB: Institute of Health Economics | Review the clinical features and outcomes of pregnant women with COVID-19. | PubMed, Web of Science, EMBASE, MEDLINE from 1 January 2020 to 16 April 2020. | 14 studies (236 COVID-19 pregnant women) | Positive CT findings (71%; 95% CI, 0.49–0.93), caesarean section (65%; 95% CI, 0.42–0.87), fever (51%; 95% CI, 0.35–0.67), lymphopenia (49%; 95% CI, 0.29–0.70), coexisting disorders (33%; 95% CI, 0.21–0.44), cough (31%; 95% CI, 0.23–0.39), fetal distress (29%; 95% CI, 0.08–0.49), preterm labor (23%; 95% CI, 0.14–0.32), severe case or death (12%; 95% CI, 0.03–0.20). Pregnant women with COVID-19 lower incidences of fever and cough. | Incidences of fever, cough and positive CT findings in pregnant women with COVID-19 are less than those in the normal population with COVID-19, rate of preterm labor is higher among pregnant women with COVID-19. No evidence of vertical transmission. |
| Goh XL 2020 | Unclear; RoB: Newcastle Ottawa Scale | To determine a more precise risk of vertical transmission, either intrauterine or during delivery. | PubMed, Medline, Embase, China National Knowledge Infrastructure until 23 May 2020. | 17 studies (402 women with COVID-19, 405 newborns) | 9/330 newborns tested positive for SARS-CoV-2. Pooled incidence of vertical transmission 16 per 1000 newborns (95% CI 3.40 to 73.11). | Risk of vertical transmission low. Quality of the evidence moderate. |

| | | | | | | |
|---|---|---|---|---|---|---|
| Han Y 2020 | No | Assess perinatal outcomes of COVID-19 infections during pregnancy and the possibility of vertical transmission. | MEDLINE, PubMed, Web of Science, Cochrane library, China National Knowledge Infrastructure (CNKI), WANFANG DATA, VIP, SinoMed, Clinical trials.gov from 1 December 2019 to 10 June 2020. | 36 studies (1103 patients) | Most common symptoms: fever (64.78%), cough (59.81%), dyspnea (23.86%). 88.73% had typical COVID-19 signs on chest CT or X-ray. Intubation in 35.87%, 4.95% mothers admitted to ICU, maternal death rate <0.01%, premature delivery 25.32%. Rate of the birth weight <2,500 g 30.65%, and that of NICU admission 24.41%. Positive nasopharynx swabs or sputum from newborns was <0.01%. | Pregnant patients with COVID-19 most commonly presented with fever, cough, dyspnea, most had positive imaging. Risk of intubation and admission to ICU were high. Risk of premature delivery was higher, leading to a high risk of NICU admission and low neonatal birthweight. Vertical transmission unlikely. |
| Hessami K 2020 | No | Review evidence on maternal, fetal and neonatal mortality cases associated with COVID-19. | PubMed, Scopus, Google Scholar, Web of Science up to 20 July 2020 | 10 studies (37 maternal and 12 perinatal mortality cases) | All maternal deaths in women with previous co-morbidities, of which the most common were obesity, diabetes, asthma and advanced maternal age. Causes of mortality: Acute respiratory distress syndrome (ARDS) and severe pneumonia, one thromboembolism. Fetal and neonatal mortalities: result severity of maternal infection or the prematurity. No evidence of vertical transmission or positive COVID-19 among neonates. | Current available evidence suggests maternal mortality mostly happened among women with previous co-morbidities and neonatal mortality seems to be a result of prematurity rather than infection. |
| Huntley BJF 2020 | Yes; RoB: Case report/case series (Murad et al.) | Review frequency of maternal and neonatal complications, as well as maternal disease severity, in SARS-CoV-2 pregnancies | MEDLINE, Ovid, ClinicalTrials.gov, MedRxiv, Scopus up to 29 April 2020. | 13 studies (538 SARS-CoV-2 pregnancies, 435 deliveries) | Maternal ICU admission 3.0% (8/263, 95% CI 1.6–5.9), maternal critical disease 1.4% (3/209, 95% CI 0.5–4.1). No maternal deaths. Preterm birth rate 20.1% (57/284, 95% CI 15.8–25.1), cesarean delivery 84.7% (332/392, 95% CI 80.8–87.9), no vertical transmission, neonatal death rate 0.3% (1/313, 95% CI 0.1–1.8). | Early pandemic data indicates low rates of maternal and neonatal mortality and no vertical transmission with SARS-CoV-2. Preterm birth rate of 20% and cesarean delivery rate >80% seems related to geographic practice patterns. |
| Juan J 2020 | Yes; RoB: Joanna Briggs Institute | Review literature on pregnancies affected by COVID-19 to evaluate the effects of COVID-19 on maternal, perinatal and neonatal outcomes. | PubMed, EMBASE, Cochrane library, China National Knowledge Infrastructure Database, Wan Fang Data up to 20 April 2020 | 19 studies (266 pregnant women with COVID-19) | Most common symptoms: fever, cough, dyspnea, fatigue. Rate of severe pneumonia low. Most had positive CT chest. 177 cases delivered, majority by Caesarean section. GA at delivery ranged from 28 to 41 weeks. Over one-third of neonates transferred to NICU. One case each of neonatal asphyxia and death. 113 neonates tested for SARS-CoV-2 were negative. 2 maternal deaths. | The clinical characteristics of pregnant women with COVID-19 are similar to those of nonpregnant adults with COVID-19. Vertical transmission unclear. |

| | | | | | | |
|---|---|---|---|---|---|---|
| Khalil A 2020 | Yes; RoB: Newcastle Ottawa Scale | Review SARS-CoV-2 infection and pregnancy, summarising evidence on the clinical features, laboratory and radiological findings, as well as the pregnancy and neonatal outcomes. | Medline, Embase, Clinicaltrials.gov, Cochrane Library up to 8 June 2020. | 86 studies (2567 COVID-19 pregnancies). | Delivery by caesarean section 48.3%. Most commonly reported symptoms: fever (63.3%), cough (71.4%), dyspnoea (34.4%). Laboratory abnormalities: raised CRP or procalcitonin (54.0%), lymphopenia (34.2%), elevated transaminases (16.0%). Preterm birth before 37 weeks' gestation 21.8%, medically-indicated (18.4%). Maternal ICU 7.0%, intubation 3.4%. Maternal mortality ~1%. Maternal ICU admission higher in cohorts with higher rates of co-morbidities (beta=0.007, p<0.05) and maternal age over 35 years (beta=0.007, p<0.01). Maternal mortality higher in cohorts with higher rates of antiviral drug use (beta=0.03, p<0.001), likely due to confounding. Neonatal NP swab positive in 1.4%. | The risk of preterm birth and caesarean delivery was increased. Maternal morbidity is similar to that of women of reproductive age. Vertical transmission probably occurs, but in small proportion of cases. |
| Kotlyar AM 2020 | Yes; RoB: Newcastle Ottawa Scale | Review current literature to determine estimates of vertical transmission of COVID-19 | PubMed, EMBASE, medRxiv, bioRxiv up to 28 May 2020. | 68 studies (979 neonates) | 27/936 SARS-CoV-2 positive neonates from mothers with COVID-19, pooled proportion of 3.2% (95% confidence interval, 2.2-4.3) for vertical transmission. SARS-CoV-2 positive samples: neonatal cord blood 2.9% (1/34), placenta samples 7.7% (2/26), amniotic fluid 0% (0/51), urine 0% (0/17), fecal or rectal swabs 9.7% (3/31). Neonatal serology positive in 3 of 82 samples (3.7%) (IgM). | Vertical transmission of SARS-Co-V-2 is possible and seems to occur in a minority of cases of maternal COVID19 infection in the third trimester. Rates of infection are similar to those of other pathogens that cause congenital infections. No data on rates of vertical transmission in early pregnancy and potential risk for consequent fetal morbidity and mortality. |
| Mahyuddin AP 2020 | No | Assess tools used to confirm maternal-fetal infection and known protective mechanisms of the placental barrier that prevent transplacental pathogen migration. | No search strategy reported. No search date reported. | 40 studies | Lack of consensus on diagnostic strategy for congenital infection for COVID-19 pregnancies. Samples positive for SARS-CoV-2: vaginal secretions (22.5%), amniotic fluid (35%), breast milk (22.5%). Neonatal COVID-19 reports: 8 studies. Histology: sparse viral particles, vascular malperfusion, placenta inflammation | Provides overview biology of maternal-fetal interface, mechanisms for vertical viral transmission. Summarises diagnostics in pregnancy to assess mother-to-child transmission. |

| | | | | | | |
|---|---|---|---|---|---|---|
| Novoa RH 2020 | Yes; RoB: Newcastle Ottawa Scale | Describe maternal clinical characteristics, maternal and perinatal outcomes in COVID-19-positive pregnant women. | MEDLINE, EMBASE, Cochrane Library and LILACS; China National Knowledge Infrastructure Database (CNKI), Chinese Science, Technology Periodical Database (VIP) and Wan Fang Data from 1 December 2019 to 27 April 2020 | 37 studies (322 infected pregnant women) | Maternal comorbidity: obesity (24.2%). 42 (28.4%) asymptomatic at admission. Symptoms: Cough (n = 148,59.7%), fever (n = 147,59.3%). Fever (OR: 0.13% CI 0.05- 0.36) and cough (0.26% CI 0.11-0.59) lower in pregnant women with COVID-19 than non-pregnant women with COVID-19. Cesarean: 99 (50.8%), vaginal delivery: 64 (32.8%). Main adverse obstetric outcome: premature birth (n = 37,18.9%). ICU: 30 (10.3%), deaths: 1 (0.3%). SARS-CoV-2 absent in breast milk, amniotic fluid, placenta, umbilical cord blood. | Pregnant women with COVID-19 less symptomatic than general population. Usual symptoms are fever and cough. Severe morbidity and mortality similar to those reported for non-pregnant women. Main adverse obstetric outcome is preterm birth. Vertical transmission unlikely in third trimester. |
| Pettirosso E 2020 | No | Describe understanding of COVID-19 illness in pregnant women, obstetric outcomes and identify gaps in knowledge. | Medline Ovid, EMBASE, WHO COVID-19 research database, Cochrane COVID-19 in pregnancy spreadsheet up to 23 May 2020. | 60 studies (1287 SARS-CoV-2 positive pregnant cases) | 8 maternal deaths, 6 neonatal deaths, 7 stillbirths, 5 miscarriages reported. 19 neonates SARS-CoV-2 positive, confirmed by RT-PCR of NP swabs. | SARS-CoV-2 infection in pregnancy often asymptomatic. Severe and critical disease similar to general population. Vertical transmission possible; unclear whether SARS-CoV-2 positive neonates infected in utero, intrapartum or postpartum. |
| Raschetti R 2020 | Yes; RoB: Mayo EBP tool | Clarify transmission route, clinical features, outcomes of COVID-19 infections. | PubMed, The Cochrane Library, Web of Science, BioXRiv, MedXRiv from 1 December 2019 to 30 August 2020 | 74 studies (176 cases of neonatal SARS-CoV-2 infections) | 70% infections due to environmental transmission, 30% vertical. 55% of infected neonates developed COVID-19; symptoms: fever (44%), gastrointestinal (36%), respiratory (52%), neurological (18%), lung imaging abnormal (64%). A lack of mother–neonate separation from birth associated with late SARS-CoV-2 infection (OR 4.94 (95% CI: 1.98–13.08), $p = 0.0002$; adjusted OR 6.6 (95% CI: 2.6–16), $p < 0.0001$), breastfeeding not associated (OR 0.35 (95% CI: 0.09–1.18), $p = 0.10$; adjusted OR 2.2 (95% CI: 0.7–6.5), $p = 0.148$). | Neonatal infections mainly occur postnatally through environmental exposure, 30% of infections may be vertical transmission.50% infected neonates develop clinical COVID-19, febrile features similar to older patients, favorable outcomes. Mother-neonate rooming-in associated with higher incidence of SARS-CoV-2 infections after 72 h of life. |

| | | | | | | |
|---|---|---|---|---|---|---|
| Rodrigues C 2020 | No | Review knowledge on impact of COVID-19 on pregnancy, describe outcome of published cases of pregnant women diagnosed with COVID-19. | PubMed, Scopus, Web of Science, MedRxiv up to 26th June 2020 | 161 studies (3,985 pregnant women with COVID-19) | 2,059 cases with pregnancy outcomes: 42 abortions, 21 stillbirths, 2,015 live births. Preterm birth: 23%. Maternal ICU: 6%, deaths: 28. Neonatal deaths: 10. From the 10/163 samples (amniotic fluid, placenta, cord blood) positive for SARS-CoV-2. 61 newborns positive for SARS-CoV-2. 4/92 breast milk samples positive. | Evidence suggests vertical transmission is possible, limited number of reported cases with intrapartum samples. Information, counseling and adequate monitoring essential to prevent and manage adverse effects of SARS-CoV-2 in pregnancy. |
| Romeo G 2020 | No | Teview case reports of MERS-CoV, SARS-CoV, SARS-CoV-2, during pregnancy and summarise clinical presentation, course of illness, pregnancy and neonatal outcomes. | MEDLINE, ClinicalTrials.gov up to 23 April 2020. | 46 studies. MERS-CoV: 8 (12 cases), SARS-CoV: 7 (17 cases), SARS-CoV-2: 31 (98 cases) | Case fatality for SARS-CoV-2 infection in pregnant women 1%. Mother-to-child transmission of MERS-CoV or SARS-CoV not observed. SARS-CoV-2 RNA detected in 7 newborns; IgM, 1 newborn. Amniotic fluid: 1 positive for SARS-CoV-2 RNA. | Limited case reports suggest possible vertical transmission of SARS-CoV-2. AR, RR, clinical significance, route of transmission unclear. Consistent data collection on timing of exposure, symptom onset, clinical presentation, course of illness, pregnancy and neonatal outcomes, and laboratory results among pregnant women with SARS-CoV-2 required. |
| Salem D 2020 | No | Review evidence on outcomes of COVID-19 infections during pregnancy. | Medline, Google Scholar from January to August 2020 | Not reported | COVID-19-positive pregnant women usually asymptomatic or mild-to-moderately symptomatic, similar to non-pregnant women. Most common clinical outcome: pneumonia. Unclear if SARS-CoV-2 infection increases risk of maternal, fetal, neonatal complications. Increased risks of complications in pregnant women with co-morbidities. Vertical transmission is possible. | Should monitor pregnant women before and after delivery, and their infants, during pandemic. |
| Sampieri CL 2020 | Yes; RoB: GRADE | Review studies on SARS-CoV-2 in clinical samples of amniotic fluid, placenta or membranes, umbilical cord blood, and human milk, from women with clinical or confirmed diagnosis of COVID-19. | PubMed from 27 March 2020 to 21 May 2020 | 17 studies (143 clinical samples: 38 amniotic fluid; 34 placentas/ membranes; 39 umbilical cord blood; 32 milk) | 9/143 samples positive for SARS-CoV-2 RNA (1 amniotic fluid before rupturing the membranes; 6 placenta/ membranes, possibility of contamination by maternal blood in 3; 2 human milk). | No studies demonstrate SARS-CoV-2 detection with viral isolation and evaluation of infective capacity of viral particles, in clinical samples of amniotic fluid, placenta/ membranes, umbilical cord blood, human milk, from women with confirmed or clinical diagnosis of COVID-19. Vertical transmission cannot be ruled out. |

| | | | | | | |
|---|---|---|---|---|---|---|
| Sheth S 2020 | No | Investigate outcomes in COVID-19 positive neonates and incidence of vertical transmission of the virus. | Pubmed, GoogleScholar from 15 November 2019 to 18 June 2020 | 39 studies (326 COVID-19 positive mothers with neonatal outcomes). | 23 neonates COVID-19 positive. Male neonates affected more (79%). 3% neonates acquired infection through suspected vertical transmission. Strict perinatal infection prevention measures can reduce horizontal transmission. Neonates asymptomatic or mildly symptomatic. No neonatal deaths. | Prognosis of COVID-19 positive neonates is good with no mortality, mild symptoms. Minimal vertical transmission. Possibility of vertical transmission very low. |
| Shrestha R 2020 | No | Summarise evidence on foetal and neonatal outcomes of pregnant women with confirmed COVID-19. | PubMed, MedRxiv from December 2019 to April 2020 | 21 studies (230 pregnant women) | Fever and cough the most common symptoms in pregnant women. Cesarean section: 162 (70.4%); 68 (29.6%) preterm deliveries. 8/161 newborns positive for SARS-CoV-2. coronavirus infection. 2 miscarriages, 2 still births, 1 neonatal death. | Outcome of pregnancy with COVID-19 in late trimester appears favourable. Preterm delivery and cesarean section higher among infected pregnant women. No conclusive evidence of vertical transmission. |
| Thomas P 2020 | Yes; RoB: CLARITY, McMaster; Joanna Briggs Institute | Summarize evidence on vertical transmission of COVID-19 infection in the third trimester and its effects on neonate. | OVID MEDLINE, EMBASE, Cochrane Central Register of Controlled Trial (CENTRAL) from January 2020 to May 2020 | 18 studies (157 mothers,160 neonates). | 5/81 (6%) neonates positive for SARS-CoV-2. Earliest test was at 16 h after birth; 1 neonate initially negative at birth, positive when re-tested, suggesting SARS-CoV-2 likely hospital-acquired rather than vertically transmitted. 13 (8%) neonates had complications or symptoms. | Rapid descriptive review; early clinical evidence suggests vertical transmission of SARS-CoV-2 from mother to neonate/newborn did not occur. |
| Tripella G 2020 | Yes; RoB: Joanna Briggs Institute | Review the impact of COVID-19 on pregnant women and neonates. | MEDLINE, EMBASE from 1 December 2019 to 18 April 2020 | 37 studies (275 pregnant women with COVID-19, 248 neonates) | Most women mild to moderate symptoms; ICU: 10; deaths: 1. 2 stillbirths; prematurity 28%. 16 neonates positive for SARS-CoV-2, 9 born from mothers infected during pregnancy. All infected neonates recovered. RT-PCR for SARS-CoV-2 negative on amniotic fluid, vaginal/cervical fluids, placenta tissue, breast milk samples. | SARS-CoV-2 infection in pregnant women associated with mild or moderate disease in most cases, low morbidity and mortality rate. Outcomes of neonates mainly favorable, although neonates at risk should be closely monitored. |

| | | | | | | |
|---|---|---|---|---|---|---|
| Turan O 2020 | Yes; RoB: NIH tools | Summarize clinical characteristics and outcomes among pregnant women hospitalized with COVID-19. | PubMed, Ovid Medline, Web of Science, China Academic Literature Database up to 29 May 2020. | 63 studies (637 women with confirmed SARS-CoV-2) | Majority (76.5%) had mild disease. Maternal fatality, 1.6%; stillbirth, 1.4%; neonatal fatality, 1.0%. Older age, obesity, diabetes mellitus, raised serum D-dimer and IL-6 predictive of poor outcomes. Pre-term births: 33.7% of which 50% were iatrogenic in women with mild COVID-19 and no complications. Most had cesarean without clear indication. 8 (2.0%) neonates positive NP test, chest infection within 48h. | Advanced gestation, maternal age, obesity, diabetes mellitus, combination of elevated D-dimer and IL-6 predictive of poor pregnancy outcomes in COVID-19. Tate of iatrogenic preterm birth and cesarean delivery high; vertical transmission possible but not proven. |
| Walker KF 2020 | No | Estimate the risk of neonate becoming infected with SARS-CoV-2 by mode of delivery, type of infant feeding and mother-infant interaction. | MEDLINE, Embase, Maternity and Infant Care Database from September 2019 to June 2020 | 49 studies (655 women, 666 neonates) | 28/666 (4%) neonates tested positive postnatally. Babies born vaginally, 8/292 (2.7%) tested positive, compared with 20/374 (5.3%) born by Caesarean. Information on feeding and baby separation lacking. 7/148 (4.7%) breastfed babies tested positive compared with 3/56 (5.3%) for formula fed. 4/107 (3.7%) babies nursed with mother tested positive, compared with 6/46 (13%) who were isolated. | Neonatal COVID-19 infection is uncommon, rarely symptomatic, and the rate of infection is no greater when the baby is born vaginally, breastfed or remains with the mother. |
| Yoon SH 2020 | No | Evaluate clinical manifestations and outcomes of neonates born to women who had COVID-19 during pregnancy. | Search databases PubMed, Embase up to 15 April 2020 | 28 studies (223 pregnant women, 201 infants) | 4 newborns born to mothers with COVID-19 tested SARS-CoV-2 positive within 48h. Breast milk, placenta, amniotic fluids, cord blood, maternal vaginal secretions all negative. Fetal death: 2; premature birth 48/185. low birth weight (< 2,500 g): 8.3%; small for gestational age: 15.6%. Birth asphyxia: 1/8%; respiratory distress syndrome 6.4%. | Evidence suggests COVID-19 during pregnancy rarely affects fetal and neonatal mortality, but can be associated with adverse neonatal morbidities. Vertical transmission has not been observed in most cases. |

**Table 4.** Findings of studies reporting placenta analysis (n=25)

| Study ID | Study design (n) | Vertical transmission | Placenta SARS-CoV-2 test result (RT-PCR) | Histopathology and Biopsy | Whole genome sequencing |
|---|---|---|---|---|---|
| Alamar I 2020 | Case report (n=1) | 1/1 | NR | Placenta ISH positive for SARS-CoV-2. Placental histopathology showed no inflammation | Not attempted |
| Birindwa EK 2020 | Case report (n=1) | 1/1 | NR | Inflammation; thrombotic vasculopathy in the placenta | Not attempted |
| Fenizia B 2020 | Cohort (n=31) | 2/31 | Positive: placental tissue in 2 cases | Generalised immune activation profile in placentae from infected compared to uninfected mothers. | Not attempted |
| Ferraiolo A 2020 | Case report (n=1) | 0/1 | Placental swabs positive | Some placental inflammation on biopsy | Not attempted |
| Grimminck K 2020 | Case report (n=1) | 0/1 | Placental sample negative | Not attempted | Not attempted |
| Kulkarni R 2020 | Case report (n=1) | 1/1 | Placental sample positive | Not attempted | Not attempted |
| Hsu AL 2020 | Case report (n=1) | 0/1 | NR | IHC with SARS-CoV-2 nucleocapsid-specific monoclonal antibody detected SARS-CoV-2 antigens throughout the placenta, under the umbilical cord, and at the central and peripheral placenta disc in chorionic villi endothelial cells, and rarely in CK7-expressing trophoblasts. | Not attempted |
| Liu W 2020 | Retrospective case analysis (n=48) | 0/48 | Placental swabs negative | Not attempted | Not attempted |

| | | | | | |
|---|---|---|---|---|---|
| Lv Y 2020 | Case report (n=1) | 0/1 | Placental swabs negative | Not attempted | Not attempted |
| Masmejan S 2020 | Retrospective case series (n=13) | 0/13 | Placental swabs negative | Not attempted | Not attempted |
| Ogamba I 2020 | Retrospective cohort (n=25) | 0/20 | NR | Placental pathology did not detect COVID-19 pathology | Not attempted |
| Oncel MY 2020 | Cohort (n=125) | 4/120 | Placental tissue negative (n=5) | Not attempted | Not attempted |
| Palalioglu RM 2020 | Case report (n=1) | 0/1 | Placental tissue negative | Not attempted | Not attempted |
| Pulinx B 2020 | Case report (n=1 mother, 2 neonates) | 2/2 (placental and amniotic fluid) | Placental swabs positive | Chronic intervillositis and extensive intervillous fibrin depositions with ischemic necrosis of the surrounding villi. Viral localization in the placental syncytiotrophoblast cells confirmed by IHC. | Not attempted |
| Rebello CM 2020 | Case report (n=1) | 1/1 | Umbilical cord blood sample positive (placenta tissue not tested) | Not attempted | Not attempted |
| Shende P 2020 | Case report (n=1) | 1/1 in amniotic fluid, also in placental cell supernatant | placental cell supernatant positive | Immunofluorescence with mAb anti-spike protein S1 and S2 on placental sections shows diffuse viral presence in cytoplasm of syncytiotrophoblasts and some cytotrophoblasts cells | Not attempted |
| Sisman J 2020 | Case report (n=1) | 1/1 | NR | Placenta histopathology indicated positive for SARS-CoV-2 | Not attempted |

| | | | | | |
|---|---|---|---|---|---|
| Smithgall MC 2020 | Cohort (n=76 placentas) | 0/51 | NR | No histomorphic changes in placenta; no viral detection by ISH and IHC. Evidence of maternal–fetal vascular malperfusion was identified, with placentas from SARS-CoV-2-positive women being significantly more likely to show villous agglutination (P = 0.003) and subchorionic thrombi (P = 0.026) than placentas from SARS-CoV-2-negative women; however no evidence for viral cause. | Not attempted |
| Stonoga E 2020 | Case report (n=1) | 0/1 | Placenta & umbilical cord blood tested positive by PCR. | Not attempted | Not attempted |
| Tang J 2020 | Case report (n=2) | 0/2 | NR | Placental tissue showed no microscopic abnormality | Not attempted |
| Vivanti AJ 2020 | Case report (n=1) | 1/1 | placenta was positive for both SARS-CoV-2 genes (E and S); Log viral load: 11.5 | Inflammation of placenta; high IHC positive Ab staining for SARS-CoV-2 cytoplasm of peri-villous trophoblastic cells. | Not attempted |
| Von Kohorn I 2020 | Case report (n=1) | 1/1 | NR | Placenta histology - no direct evidence of virus | Not attempted |
| Zaigham M 2020 | Case report (n=1) | 1/1 | NR | high viral load in placenta with massive perivillous fibrin deposition, acute intervillositis in areas with strong positivity for SARS-CoV-2 and chorangiosis in the areas less affected by infection and inflammation | Whole genome sequencing of isolates from the mother & placenta: Presence of |

| | | | | | |
|---|---|---|---|---|---|
| | | | | | single variant of the virus |
| Zhang P 2020 | Case series (n= 364 placenta samples) | 2/53 | NR | SARS-CoV-2 detected by ISH in placenta of positive cases (2/ 53 placental samples from PCR positive mothers ). No histopathological features | Not attempted |
| Zheng T 2020 | Case report (n=2) | 0/2 | NR | Both placentas were examined and found to be normal | Not attempted |

**Table 5.** Ct and RNA concentration results from included studies.

| Study | PCR cycle methods | Ct counts | RNA concentration / viral load | Live culture attempted (positivity rate) |
|---|---|---|---|---|
| Alamar I 2020 | NR | NR | NR | Not attempted |
| Anand P 2020 | First screen using E gene, then if +ve, two genes, RdRp & ORF-1b-nsp14b. The detection limit was 35 for E, RDRp & ORF. | 35? Unclear | NR | Not attempted |
| Ayed A 2020 | NR | NR | NR | Not attempted |
| Bachani S 2020 | E, RDRp & ORF-1bnsp14b. The threshold value for infectivity </= 35 cycles for E, RDRp & ORF genes. | 35 | Lowest Cts observed for neonate 3: Ct = 15.82, DOL4; & neonate 4: Ct = 15.33, DOL1. | Not attempted |
| Bandyopadhyay T 2020 | NR | NR | NR | Not attempted |
| Barbero P 2020 | NR | NR | NR | Not attempted |

| | | | | |
|---|---|---|---|---|
| Birindwa EK 2020 | NR | NR | NR | Not attempted |
| Bordbar A 2020 | NR | NR | NR | Not attempted |
| Chaudhary S 2020 | NR | NR | NR | Not attempted |
| Cojocaru L 2020 | NR | NR | NR | Not attempted |
| Demirjian A 2020 | NR | maternal NP DOL1 Ct = 34.9; DOL3 = 39.4 | NR | Not attempted |
| Farsi Z 2020 | NR | NR | NR | Not attempted |
| Fenizia B 2020 | RdRP, N & E genes simultaneously amplified & tested. Ct-value <40 defined as positive test result according to manufacturer's instruction. | <40 for two newborns (remainder 40+ judged negative) | NR | Not attempted |
| Ferraiolo A 2020 | 3 target genes: RdRP, E & N, & a process control. | NR | NR | Not attempted |
| Gale C 2020 | NR | NR | NR | Not attempted |
| Gao W 2020 | NR | NR | NR | Not attempted |

| | | | | |
|---|---|---|---|---|
| Gao X 2020 | RT-PCR assays for SARS-CoV-2 ORF1ab/N genes | NR | NR | Not attempted |
| Grimminck K 2020 | NR | NR | NR | Not attempted |
| He Z 2020 | NR (state following WHO guidelines) | NR | NR | Not attempted |
| Hinojosa-Velasco A 2020 | real-time RT-PCR, ORF1ab & N gene targets. | 23 | reported as "high" based on Ct of 23 | Not attempted |
| Hsu AL 2020 | NR | NR | NR | Not attempted |
| Hu X 2020 | NR | NR | NR | Not attempted |
| Khan MA 2020 | NR | NR | NR | Not attempted |
| Kulkarni R 2020 | NR | NR | NR | Not attempted |
| Liu W 2020 | NR | NR | NR | Not attempted |
| Luo Q 2020 | ORF1ab gene & N gene | NR | NR | Not attempted |

| | | | | |
|---|---|---|---|---|
| Lv Y 2020 | NR | NR | NR | Not attempted |
| Maraschini A 2020 | NR | NR | NR | Not attempted |
| Marin Gabriel M 2020 | NR | NR | NR | Not attempted |
| Masmejan S 2020 | NR | NR | NR | Not attempted |
| Mohakud NK 2020 | NR | NR | NR | Not attempted |
| Moreno SC 2020 | NR (state methods according to WHO guidelines) | NR | NR | Not attempted |
| Nayak AH 2020 | NR | NR | NR | Not attempted |
| Ogamba I 2020 | NR | NR | NR | Not attempted |
| Olivini N 2020 | NR | NR | NR | Not attempted |
| Oncel MY 2020 | NR (state methods according to WHO guidelines) | NR | NR | Not attempted |

| | | | | |
|---|---|---|---|---|
| Pace RM 2020 | NR | NR | NR | Not attempted |
| Palalioglu RM 2020 | NR | NR | NR | Not attempted |
| Parsa Y 2020 | NR | NR | NR | Not attempted |
| Pereira A 2020 | NR | NR | NR | Not attempted |
| Pessoa FS 2020 | NR | NR | NR | Not attempted |
| Pissarra S 2020 | NR | NR | NR | Not attempted |
| Popofsky S 2020 | NR | NR | NR | Not attempted |
| Pulinx B 2020 | rRT-PCR based on the CDC oligonucleotide primers & probes for detection of N gene | at birth placental tissue samples Ct = 33 & Ct = 30; amniotic fluid Ct = 23; maternal blood Ct = 35) | NR | Not attempted |
| Rebello CM 2020 | NR | NR | NR | Not attempted |

| | | | | |
|---|---|---|---|---|
| Rivera-Hernandez P 2020 | NR | NR | NR | Not attempted |
| Rubio Lorente AMR 2020 | NR | NR | NR | Not attempted |
| Sajjan GR 2020 | NR | NR | NR | Not attempted |
| Schwartz DA 2020 | NR | NR | NR | Not attempted |
| Shende P 2020 | RT-PCR done as per the ICMR/WHO Guidelines for RdRP & E genes | Ct for E gene 26.3, Ct for RdRp gene 25.4 (amniotic fluid) | NR | Not attempted |
| Singh MV 2020 | NR | NR | NR | Not attempted |
| Sisman J 2020 | NR | NR | NR | Not attempted |
| Smithgall MC 2020 | NR | NR | NR | Not attempted |
| Stonoga E 2020 | N and ORF1ab; Ct value considered +ve if both viral genes are <38. | umbilical cord blood N gene Ct = 30.3, ORF1ab Ct = 31.9; placenta N gene Ct = 25.5, ORF1ab Ct = 24.5 | NR | Not attempted |

| | | | | |
|---|---|---|---|---|
| Tang F 2020 | two sets of primers to respectively amplify ORF1ab & N gene | NR | NR | Not attempted |
| Tang J 2020 | NR | NR | NR | Not attempted |
| Vendola N 2020 | NR | NR | NR | Not attempted |
| Vinuela MC 2020 | RT-PCR detection of N & ORF 1a1b genes; Ct for N gene recorded | Ranged from 36 to 41; median 40. | NR | Not attempted |
| Vivanti AJ 2020 | RT-PCR detection of E & S gene of SARS-CoV-2; full details of PCR methods provided | 40 | Viral load (log) mother: NP: 4.22; vaginal: 0.63; placenta: 11.15; amniotic: 2.09; blood: 4.87. Neonate: blood: 1.15; NP (DOL1): 2.21; rectal: 4.71 | Not attempted |
| Von Kohorn I 2020 | Two RT-PCRs: Test1: E & N2 gene; Test 2: ORF1 & E gene | For positive tests DOL2-7: range from 17.7 (DOL7) to 44.1 (DOL2) | NR | Not attempted |
| Vouga M 2020 | NR | NR | NR | Not attempted |
| Woodworth KR 2020 | NR | NR | NR | Not attempted |

| | | | | |
|---|---|---|---|---|
| Yang R 2020 | NR | NR | NR | Not attempted |
| Zaigham M 2020 | RT-PCR for envelope gene, details of PCR methods provided | Maternal: NP Ct = 26.7; blood Ct = 31.7. Infant: NP DOL 2 to 16: Ct range 18.1 (DOL5) to 36.1(DOL12). Placenta: Day of delivery Ct = 13.6 | NR | Not attempted |
| Zhang L 2020 | NR | NR | NR | Not attempted |
| Zhang P 2020 | NR | NR | NR | Not attempted |
| Zheng T 2020 | NR | NR | NR | Not attempted |
