## Supplementary material for "SARS-CoV-2 and the role of vertical transmission from infected pregnant women to their fetuses: systematic review": List of excluded studies

**List of excluded studies n=8**

No relevant data included n= 7
Study was a subset of data of an included study n=1

Barragan M 2020

Barragan M, Guillén JJ, Martin-Palomino N, Rodriguez A, Vassena R. Undetectable viral RNA in oocytes from SARS-CoV-2 positive women. Hum Reprod. 2021 Jan 25;36(2):390-394. doi: 10.1093/humrep/deaa284. PMID: 32998162; PMCID: PMC7543480.

Chambers CD 2020

Chambers CD, Krogstad P, Bertrand K, Contreras D, Tobin NH, Bode L, Aldrovandi GM. Evaluation of SARS-CoV-2 in Breastmilk from 18 Infected Women. medRxiv [Preprint]. 2020 Jun 16:2020.06.12.20127944. doi: 10.1101/2020.06.12.20127944. Update in: JAMA. 2020 Aug 19;: PMID: 32587991; PMCID: PMC7310649.

Chu H 2020

Chu H, Li J, Yan J, Bai T, Schnabl B, Zou L, Yang L, Hou X. Persistent SARS-CoV-2 RNA Positive in Feces but Negative in Breastmilk: A Case Report of COVID-19 in a Breastfeeding Patient. Front Med (Lausanne). 2020 Dec 2;7:562700. doi: 10.3389/fmed.2020.562700. PMID: 33344466; PMCID: PMC7738631.

Han T 2020

Han T. Outbreak investigation: transmission of COVID-19 started from a spa facility in a local community in Korea. Epidemiol Health. 2020;42:e2020056. doi: 10.4178/epih.e2020056. Epub 2020 Jul 29. PMID: 32777883; PMCID: PMC7871164.

Marin Gabriel 2020 Acta Paediatr

Marín Gabriel MA, Cuadrado I, Álvarez Fernández B et al. Multicentre Spanish study found no incidences of viral transmission in infants born to mothers with COVID-19. Acta Paediatr. 2020 Nov;109(11):2302-2308. doi: 10.1111/apa.15474. Epub 2020 Jul 28. PMID: 32649784; PMCID: PMC7404522. [subset of the included study Marin Gabriel 2020 The Pediatric Infectious Disease Journal (confirmed with the authors)]

Pace RM 2020

Pace RM, Williams JE, Järvinen KM et al. COVID-19 and human milk: SARS-CoV-2, antibodies, and neutralizing capacity. medRxiv [Preprint]. 2020 Sep 18:2020.09.16.20196071. doi: 10.1101/2020.09.16.20196071. PMID: 32995804; PMCID: PMC7523143.

Rubin ES 2020

Rubin ES, Sansone SA, Hirshberg A, Clement EG, Srinivas SK. Detection of COVID-19 in a Vulvar Lesion. Am J Perinatol. 2020;37(11):1183-1184. doi:10.1055/s-0040-1713665

Yu Y 2020

Yu Y, Li Y, Hu Y et al. Breastfed 13 month-old infant of a mother with COVID-19 pneumonia: a case report. Int Breastfeed J 15, 68 (2020). https://doi.org/10.1186/s13006-020-00305-9

**Duplicates in the search database of included studies n=9**
Direct duplicate n=7
Duplicate is the preprint version of an included peer reviewed publication n=2

Ayed A 2020 preprint duplicate of included peer reviewed study

Ayed A, Embaireeg A, Benawath A et al.Maternal and perinatal characteristics and outcomes of pregnancies complicated with COVID-19 in Kuwait. MedRxiv doi: https://doi.org/10.1101/2020.07.10.20150623
Now published in BMC Pregnancy and Childbirth doi: 10.1186/s12884-020-03461-2

Grimminck K 2020 duplicate of included study

Grimminck K, Santegoets LAM, Siemens FC, Fraaij PLA, Reiss IKM, Schoenmakers S. No evidence of vertical transmission of SARS-CoV-2 after induction of labour in an immune-suppressed SARS-CoV-2-positive patient. BMJ Case Rep. 2020 Jun 30;13(6):e235581. doi: 10.1136/bcr-2020-235581. PMID: 32606133; PMCID: PMC7358094.

Hsu AL 2020 MedRxiv preprint duplicate of included peer reviewed study

Hsu AL, Guan M, Johannesen E et al. Placental SARS-CoV-2 in a patient with mild COVID-19 disease. MedRxiv https://doi.org/10.1101/2020.07.11.20149344.
Now published in J Med Virol

Hu X duplicate of included study

Hu X, Gao J, Wei Y, Chen H, Sun X, Chen J, Luo X, Chen L: Managing Preterm Infants Born to COVID-19 Mothers: Evidence from a Retrospective Cohort Study in Wuhan, China. Neonatology 2020;117:592-598. doi: 10.1159/000509141

Khan MA 2020 duplicate of included study

Khan MA, Kumar V, Ali SR. Vertical Transmission of Novel Coronavirus (COVID-19) from Mother to Newborn: Experience from a Maternity Unit, The Indus Hospital, Karachi. J Coll Physicians Surg Pak. 2020 Oct;30(10):136. doi: 10.29271/jcpsp.2020.supp2.136. PMID: 33115590.

Masmejan S 2020 duplicate of included study

Masmejan S, Pomar L, Favre G, Panchaud A, Giannoni E, Greub G, Baud D. Vertical transmission and materno-fetal outcomes in 13 patients with coronavirus disease 2019. Clin Microbiol Infect. 2020 Nov;26(11):1585-1587. doi: 10.1016/j.cmi.2020.06.035. Epub 2020 Jul 8. PMID: 32652239; PMCID: PMC7341030.

Rivera-Hernandez P 2020 duplicate of included study

Rivera-Hernandez P, Nair J, Islam S, Davidson L, Chang A, Elberson V. Coronavirus Disease 2019 in a Premature Infant: Vertical Transmission and Antibody Response or Lack Thereof. AJP Rep. 2020 Jul;10(3):e224-e227. doi: 10.1055/s-0040-1715176. Epub 2020 Aug 20. PMID: 33094009; PMCID: PMC7571559.

Rubio Lorente AM 2020 duplicate of included study

Rubio Lorente AM, Pola Guillén M, López Jimenez N, Moreno-Cid Garcia-Suelto M, Rodriguez Rodriguez E, Pascual Pedreño A. Study of amniotic fluid in pregnant women infected with SARS-CoV-2 in first and second trimester. Is there evidence of vertical transmission? J Matern Fetal Neonatal Med. 2020 Aug 30:1-3. doi: 10.1080/14767058.2020.1811669. Epub ahead of print. PMID: 32862730.

Vendola N 2020 duplicate of included study

Vendola N, Stampini V, Amadori R, Gerbino M, Curatolo A, Surico D. Vertical transmission of antibodies in infants born from mothers with positive serology to COVID-19 pneumonia. Eur J Obstet Gynecol Reprod Biol. 2020 Oct;253:331-332. doi: 10.1016/j.ejogrb.2020.08.023. Epub 2020 Aug 27. PMID: 32878687; PMCID: PMC7449895
